# Nanopore sequencing enables highly accurate genotyping and identification of resistance determinants in key nosocomial pathogens

**DOI:** 10.1101/2025.07.24.25332173

**Authors:** Hugh Cottingham, Louise M. Judd, Taylor Harshegyi-Hand, Jessica A. Wisniewski, Luke V. Blakeway, Tiffany Hong, Matthew H. Parker, Anton Y. Peleg, Kathryn E. Holt, Jane Hawkey, Nenad Macesic

## Abstract

Whole genome sequencing of bacterial pathogens can positively impact infectious disease management in clinical contexts, both in individual settings and assisting infection prevention efforts. However, logistical issues have often prevented its translation into clinical settings. Oxford Nanopore Technologies (ONT) platforms are flexible, affordable, and now offer accuracy comparable to other sequencing platforms, making them uniquely well-suited for clinical bacterial isolate sequencing. We sought to determine the best methods for implementing ONT sequencing into clinical settings by benchmarking multi-locus sequence typing (MLST), core genome MLST (cgMLST), antimicrobial resistance (AMR) and core genome single nucleotide polymorphism (cgSNP) typing against the genomic gold standard. We sequenced 199 *Enterobacterales* isolates with Illumina and ONT platforms and assessed performance based on sequencing chemistry, basecaller, basecalling model, assembly status, assembly polishing and sequencing depth. Modern ONT data generated perfect MLST and AMR allelic variant calls, and correctly classified a median of 99.5% of cgMLST loci. Illumina and kmer-based SNP typing failed to call 9-68 SNPs per 1000 sites due to poor sensitivity in repetitive regions of the reference genome, while ONT’s long reads generated perfect SNP calls across the entire genome using simulated readsets. Using real sequencing data to identify putative transmission pairs, ONT read-based methods were concordant with traditional Illumina approaches in 155-158/158 (98.1-100%) of isolate pairs. We also provide specific recommendations on sequencing depth and basecalling model based on the time and computational resources available to the user. This study demonstrates the viability of modern ONT data for highly accurate characterisation of bacterial pathogens while providing an actionable framework for clinical implementation.

## Introduction

Whole genome sequencing (WGS) of human pathogens has become a crucial tool in research and public health settings (1). Despite the potential advantages of sequencing platforms such as Illumina and Oxford Nanopore Technologies (ONT) for identifying resistance mechanisms and monitoring suspected outbreaks, translation into clinical settings has been more difficult. Illumina has historically been the more popular choice in research contexts due to its higher accuracy rate (∼99.9%) compared to ONT (∼95-97%) (2, 3). However, it has not been routinely implemented into clinical settings due to a combination of high instrument costs, prohibitive consumables costs for small batch sizes and analysis limitations due to its short (max 301bp) read lengths.

ONT is affordable (USD $600-800 per flowcell) with consistent consumable costs regardless of batch size, offers real-time analysis, and generates long (20 kbp +) reads that assist in various downstream analyses. The release of the ‘super’ basecalling model and R10.4.1 flowcells has also improved accuracy to levels closer to Illumina, achieving ∼99% read accuracy and generating assemblies with less than one error per 100,000 base pairs (3, 4). ONT platforms allow users to customise various aspects of data generation and analysis, including sequencing duration, basecalling algorithms, assembly methods and characterisation software. While this flexibility is part of what makes ONT uniquely viable for clinical implementation, it is not yet clear how different combinations of sequencing depth, basecalling algorithms and assembly methods perform in genotyping and identifying resistance determinants in bacterial isolates.

Previous benchmarking has found modern R10.4.1 ONT data to be comparable to Illumina in MLST, cgMLST, AMR and cgSNP-based typing of bacterial pathogens (5–14). However, these studies did not adequately assess the performance of i) using different basecalling software and models; ii) using raw reads versus assemblies as input; iii) characterising allelic variants of clinically important AMR genes. Specifically, to our knowledge there has been no attempt to translate the results of these benchmarking analyses into defined recommendations for clinical sequencing of bacterial isolates.

In this study we addressed this gap by investigating the effects of flowcell pore type, basecalling model, sequencing depth, assembly vs raw reads and assembly polishing on MLST, cgMLST, AMR gene detection and core genome single nucleotide polymorphism (cgSNP)-based analyses. This involved sequencing 199 bacterial *Enterobacterales* isolates and comparing ONT-only results to hybrid assembly genomic gold standards. We also provide recommendations for the best combination of sequencing and analysis methods depending on the time and computational resources of the user. This work provides an important roadmap for use of modern ONT sequencing as a frontline surveillance and diagnostic technology in clinical settings.

## Results

### Benchmarking Dataset

We generated input datasets from 199 bacterial isolates that varied according to sequencing chemistry, sequencing depth, basecaller, basecalling model, assembly status and assembly polishing (**Figure 1**, **Table 1, Supp. Table 1, Supp. Figure 1**, see **Methods**). We then generated MLST, cgMLST, AMR and cgSNP calls using read and assembly-based tools and assessed accuracy by comparing to gold standard hybrid assemblies of the same strain (**Figure 1**, see **Methods**).

**Figure 1.**
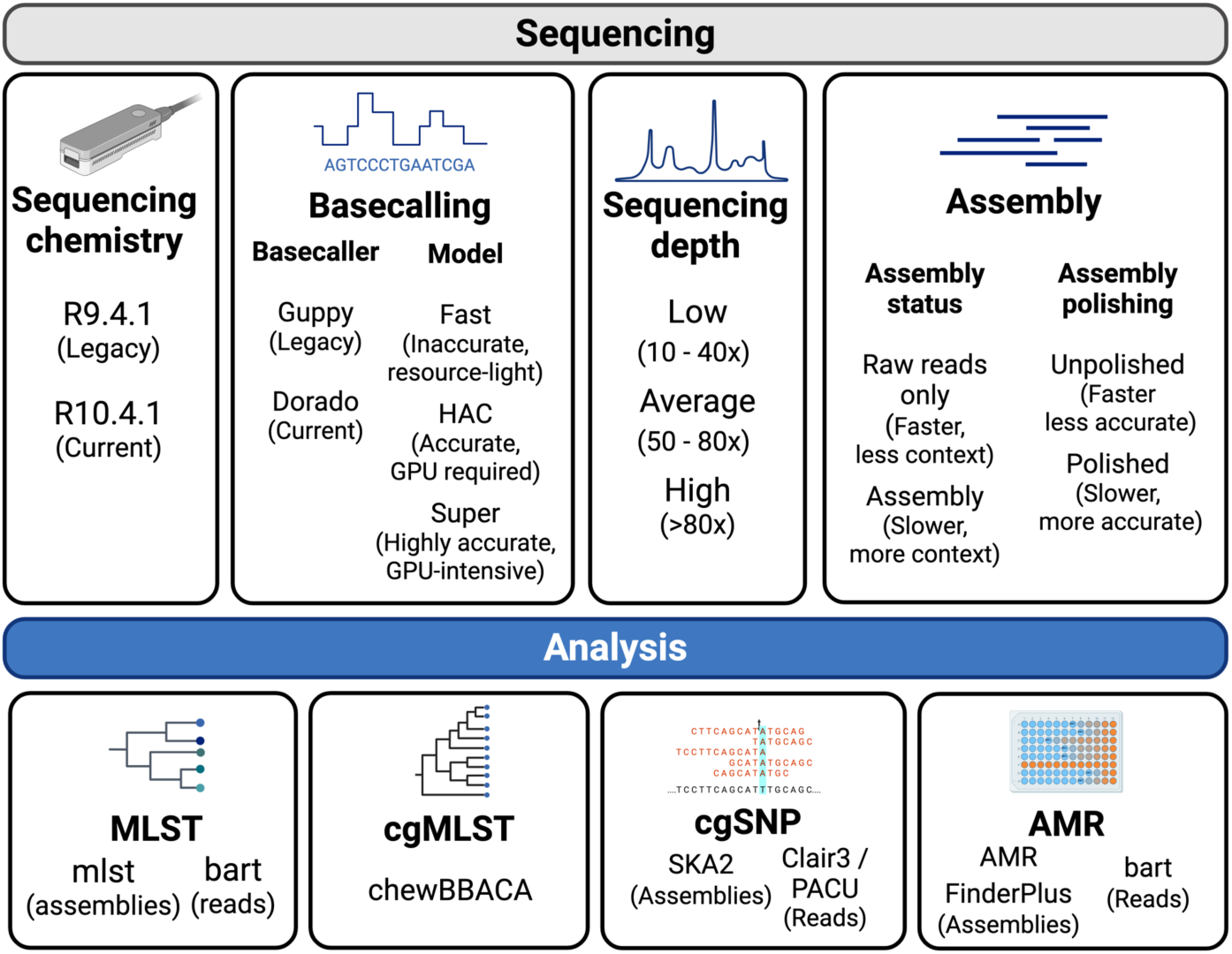
– Overview of analysis methods used to generate an ONT-only dataset to benchmark MLST, cgMLST, cgSNP and AMR typing performance.

**Table 1.** – Summary statistics for isolates included in benchmarking analyses.

| Species group | Total isolates |  | Unique STs |  | Prominent STs (n=no. isolates) |  |
| --- | --- | --- | --- | --- | --- | --- |
|  | R9.4.1 | R10.4.1 | R9.4.1 | R10.4.1 | R9.4.1 | R10.4.1 |
| *K. pneumoniae*<br>species complex | 60 | 25 | 39 | 9 | ST37 (n=4)<br>ST323 (n=5) | ST29 (n=10)<br>ST323 (n=9) |
| *E. coli* | 27 | 33 | 16 | 10 | ST10 (n=3)<br>ST38 (n=4)<br>ST617 (n=3) | ST95 (n=14)<br>ST131 (n=12) |
| *E. cloacae*<br>complex | 27 | 27 | 9 | 4 | ST93 (n=7)<br>ST114 (n=7)<br>ST190 (n=9) | ST93 (n=13)<br>ST190 (n=13) |

### Modern ONT data generated perfect MLST calls across tested species

Basecalling model was the most important factor for MLST accuracy: High accuracy (HAC) and Super (SUP) assemblies generated a correct call in mean 96.1% and 98.8% of isolates, respectively, across all species, basecallers and chemistries, compared to the Fast model achieving this in only 41.7% of isolates (**Supp. Figure 2, Supp. Table 2**). Sequencing chemistry also impacted accuracy: R10.4.1 sequences basecalled with Dorado generated perfect MLST calls in every isolate using both HAC and SUP models. However, using legacy R9.4.1 SUP data led to four incorrect calls in *K. pneumoniae* species complex (KpSC) species, while errors in all three species groups were noted when R9.4.1 HAC data were used.

**Table 2.**
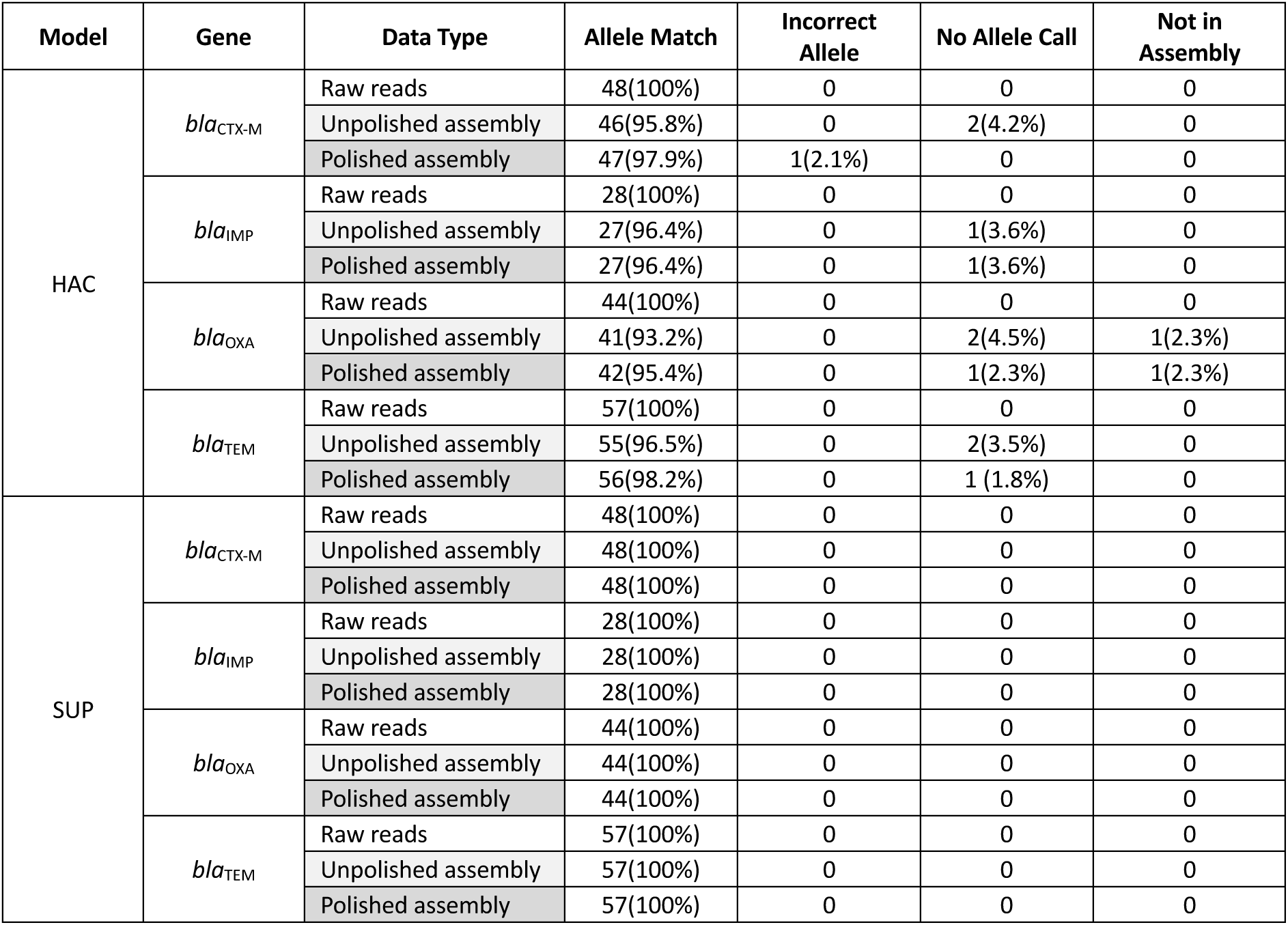
– Accuracy of AMR allelic variants using Dorado R10.4.1 raw reads, unpolished assemblies and polished assemblies. Maximal depth (100x and below) was used for each isolate. No Allele Call refers to instances where AMRFinderPlus detected the gene but could not provide a confident allele call.

We then assessed performance of modern ONT data (R10.4.1, Dorado, HAC or SUP models) when unpolished assemblies, polished assemblies and raw reads were used (**Figure 2A, Supp. Table 3**). We noted that assembly polishing was not necessary, with both unpolished and Medaka-polished assemblies generating 100% MLST calls across all R10.4.1 isolates using both HAC and SUP. Read-based MLST performed almost as well as assemblies, with only one incorrect locus in an *E. coli* isolate (*icd* locus in AH20I049) across the entire dataset. This appeared to be an error generated by the read-based tool bart (15), which aligns reads to the MLST database using KMA (16) and parses its output. Running KMA by itself against the candidate alleles at this locus generated a higher alignment score for the correct allele, suggesting that the error was made by bart rather than the ONT basecaller Dorado.

**Figure 2.**
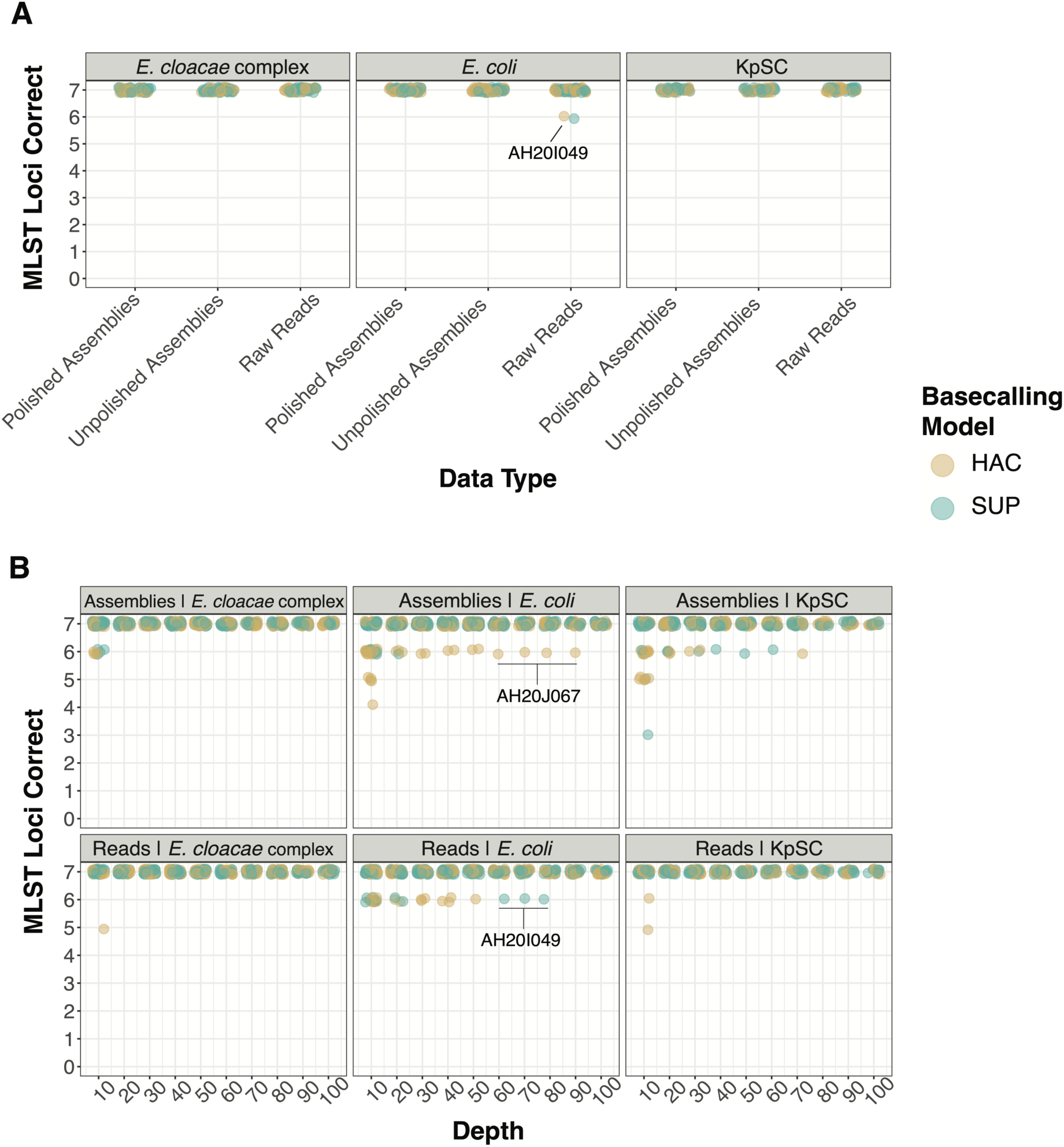
– Accuracy of MLST genotyping using. **A)** raw reads, unpolished assemblies and polished assemblies at maximum available depth (100x or below) for each isolate and **B)** raw reads and polished assemblies at depths from 10x – 100x. Labelled *E. coli* isolates (AH20J067 and AH20I049) refer to strains that had incorrect calls at multiple depths. KpSC strains are not labelled as errors came from a variety of strains.

To assess the effects of sequencing depth and basecalling model, we generated MLST calls using subsets of each readset up to 100x depth (see **Methods**). Raw reads generated comparable or better calls compared with assembly methods for *E. cloacae* complex (ECC) and KpSC, generating 100% MLST calls in ECC and KpSC at all depths greater than 10x using both HAC and SUP (**Figure 2B**). For *E. coli,* SUP assemblies outperformed raw reads, generating perfect calls from 30x and above. KpSC and *E. coli* assembly errors at submaximal depths were predominantly due to adenine insertion errors in small homopolymer regions of MLST allele-encoding regions. In KpSC, this occurred in both HAC and SUP assemblies, while in *E. coli* it only occurred in HAC assemblies.

### Modern ONT data reliably classified cgMLST loci in *E. coli* and KpSC isolates

While MLST is a traditional strain typing method, it lacks resolution when dealing with highly diverse STs or with species that have a poorly defined MLST scheme. In these cases, cgMLST can be used to differentiate strains based on allelic profiles across thousands of genetic loci, as opposed to a limited number used in MLST (17). We generated cgMLST calls in *E. coli* and KpSC subset assemblies and compared them to the gold standard (see **Methods**). *E. cloacae* does not have an established cgMLST scheme, so was not included in this analysis.

Using our optimal approach (R10.4.1 chemistry, maximum available depth for each strain, Dorado SUP basecalling and Medaka polishing), both species had a median of 11 incorrect loci (0.5% of total loci) per isolate (**Supp. Table 4**). For *E. coli,* most errors came in one strain (ECamp083) that had incorrect allelic calls in 118 loci (**Supp. Figure 3A**), or six loci (*b0197, b0241, b1704, b2172, b4354, b1819*) where most strains did not generate any allele call (**Supp. Figure 3B**). We found exact matches between ONT-only and hybrid assemblies in these six genes following alignment with Minimap2 (18), suggesting that errors likely arose during CDS prediction by the cgMLST tool (chewBBACA (19)). Sequencing depth (47x) for ECamp083 was amongst the lowest of all isolates and likely contributed to the higher error rate. For KpSC, the overall random error rate was higher, although there was no particularly error-prone strain comparable to ECamp083 in *E. coli* (**Supp. Figure 3A**). Allele calls were not generated for two loci (*KP1_RS00725* and *KP1_RS25355*), similar to the issues detailed for six loci in *E. coli* (**Supp. Figure 3B**).

When we evaluated the impact of basecalling model, we found that using SUP basecalling provided a higher increase in performance over HAC compared to MLST typing (median 11 incorrect cgMLST loci per isolate for both species using SUP, median 22 and 29 incorrect for *E. coli* and KpSC, respectively, using HAC) (**Supp. Table 4**). We also found that accuracy improved with increasing depth before plateauing above 50x, similar to the results of MLST (**Supp. Figure 4**).

A common downstream analysis after calling cgMLST alleles is to group strains into genetic clusters based on the number of genes by which they differ. For *E. coli* and KpSC, clusters are often defined as strains that have ten or less gene differences across all cgMLST loci (20). In our dataset, there were no *E. coli* strains that shared a cluster, so we focused on KpSC.

When applying this clustering threshold of ten allele differences to the optimal ONT-only KpSC assemblies (R10.4.1, Dorado, SUP, Medaka-polished, max depth), we found that most clusters were not effectively resolved when compared to the gold standard assemblies (**Figure 3**). As shown in **Supp. Figure 3A**, several ONT-only assemblies differed to their gold standard assembly by up to 40 loci, indicating that a clustering threshold of ten alleles may be too sensitive. We then evaluated a clustering threshold of 50 alleles. We found that cgMLST clusters 1, 3 and 4 contained the same isolates as the gold standard. Meanwhile, cluster 2 still contained both false positive and false negative errors (**Figure 3**).

**Figure 3.**
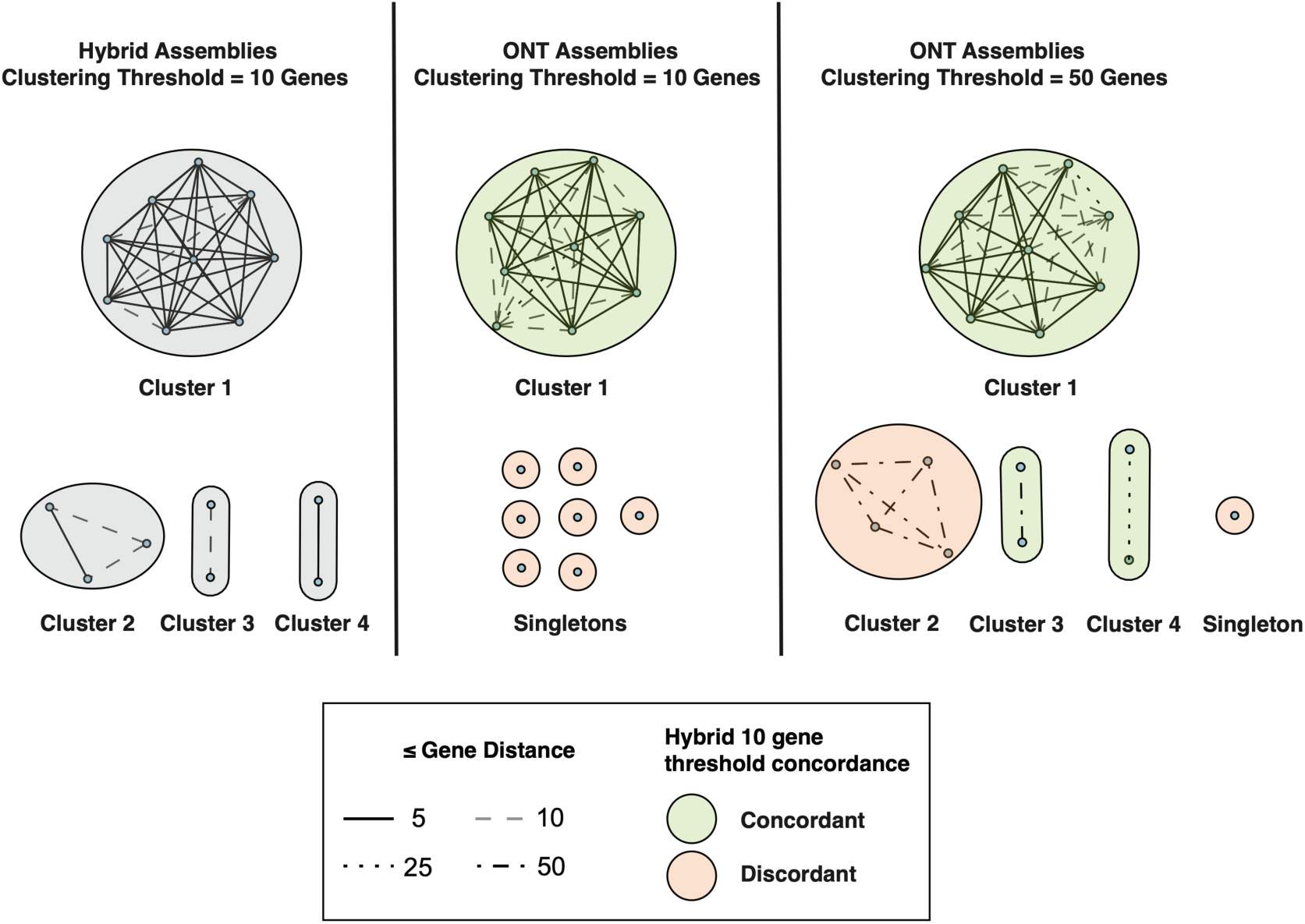
– Hierarchical clustering of KpSC hybrid and ONT-only assemblies based on varying cgMLST distance thresholds. Each node represents an isolate. Clusters with two or more strains are numbered. Gene distances within clusters are depicted as varied line types depending on distance value. Singletons are isolates that clustered using the hybrid assembly 10 gene threshold but did not cluster when using ONT assemblies.

When comparing modern ONT data to legacy datasets in cgMLST typing accuracy, R10.4.1 data (median 98.65% of loci correct) far outperformed R9.4.1 (median of 93.9% of loci correct) using HAC and SUP models (**Supp. Table 5**). Fast basecalling again performed poorly across R9.4.1 and R10.4.1 datasets (median 53.3% of loci correct).

### Modern ONT data led to 100% accurate typing of allelic variants in acquired AMR genes

To assess AMR allelic variant calling in our benchmarking dataset, we analysed all isolates for which the gold standard assembly contained the clinically important *bla*_CTX-M,_ *bla*_IMP,_ *bla*_OXA_ or *bla*_TEM_ acquired beta-lactamase genes. We ran assembly-based and read-based AMR detection tools on ONT-only datasets and compared the allele calls to that of the gold standard genome (see **Methods**). R10.4.1 SUP datasets generated 100% allelic matches across all four genes using both raw reads and assemblies (**Table 2, Supp. Table 6**). Raw HAC reads outperformed HAC assemblies, generating 100% allelic matches compared to 95.5% in unpolished assemblies and 96.8% in polished assemblies. Raw reads also excelled at lower depths, generating 100% allelic matches from 30x and above across all four genes (**Supp. Figure 5**). SUP assemblies reached 100% accuracy at 50x and above, while HAC showed 1-2 errors in at least one gene at all depths except 100x.

When comparing modern ONT data to legacy datasets, we found that R10.4.1 HAC assemblies showed similar accuracy (96.8%) to R9.4.1 SUP assemblies (95.5%) (**Supp. Figure 6, Supp. Table 7**). The Fast basecalling model performed worse than HAC and SUP for all datasets, generating a correct allele call in an average of just 62.6% of cases across all genes and chemistries compared to 96.3 and 97.7% for HAC and SUP respectively (**Supp. Figure 6, Supp. Table 7**). R9.4.1 Fast assemblies (84.4% allelic match) outperformed R10.4.1 Fast assemblies (40.8%), although this was likely due to there being no Medaka polishing model for Dorado Fast data types.

While antimicrobial activity of these beta-lactamase genes is often dependent on allelic variation, for many other AMR genes resistance phenotype can be predicted from gene presence. We evaluated AMR gene detection in R10.4.1 assemblies at depths up to 100x (see **Methods**). We noted that structural errors in ONT-only assemblies led to omission of AMR genes entirely, particularly at lower depths (**Supp. Figure 7**). Omission rates across all genes with low-depth assemblies (40x and below) were 9.2%, 6.2% and 7.4% for Fast, HAC and SUP, respectively. Medium-high depth assemblies (50x and above) still had missing AMR genes in some cases, although at a lower rate of 1.2% across all genes and models. These findings also appeared to be gene-specific, with tetA, blaOXA, and blaCTX-M showing omission rates of 7.1%, 1.5% and 3.1% even at 50x and above. Location of AMR genes also impacted omission rates: plasmid-located genes had a mean 1.5% omission rate across all genes versus 0.07% for chromosomally-located genes at depths of 50x and above.

### ONT read-based variant callers outperformed Illumina reads and ONT assemblies using simulated data

One of the limitations of legacy ONT sequencing was concerns surrounding read accuracy, making its use in analyses such as variant calling impractical. Read accuracy of modern ONT data has now risen to ∼99% (3), but further data is needed to confirm that this leads to more accurate variant calling. To gain an understanding of how variant callers performed under highly controlled conditions with modern ONT data, we benchmarked four variant calling tools on simulated R10.4.1 Dorado SUP readsets generated from *E. cloacae, E. coli* and *K. pneumoniae* genomes with known SNP sites (see **Methods**). ONT read-mapping tools (Clair3, PACU) outperformed ONT assembly (SKA2) and Illumina read-mapping (Reddog) at 40x, 70x and 100x, generating perfect SNP profiles at 100x compared to 9-22 and 14-29 errors per 1000 SNPs for Reddog and SKA2, respectively (**Figure 4**). Reddog and SKA2 had an elevated false negative rate (**Figure 4A**) caused by an inability to resolve repeat regions longer than the read or max kmer length for Reddog and SKA2, respectively (see **Methods** for description of defining repetitive regions). Clair3 and PACU utilised ONT’s long reads to accurately call SNPs in these regions, and at 100x depth made no errors. Nearly all false positive errors (**Figure 4B**) came from SKA2. These sites were heterozygous for the alternative and reference allele, which can be identified using mapping approaches but not from standard assembly methods. Incorrect SNP allele calls (**Figure 4C**) were mostly generated by Clair3 and PACU at lower sequencing depths – this is indicative of ONT’s slightly higher raw read error rate compared to Illumina reads (Reddog) and ONT consensus accuracy (SKA2).

**Figure 4.**
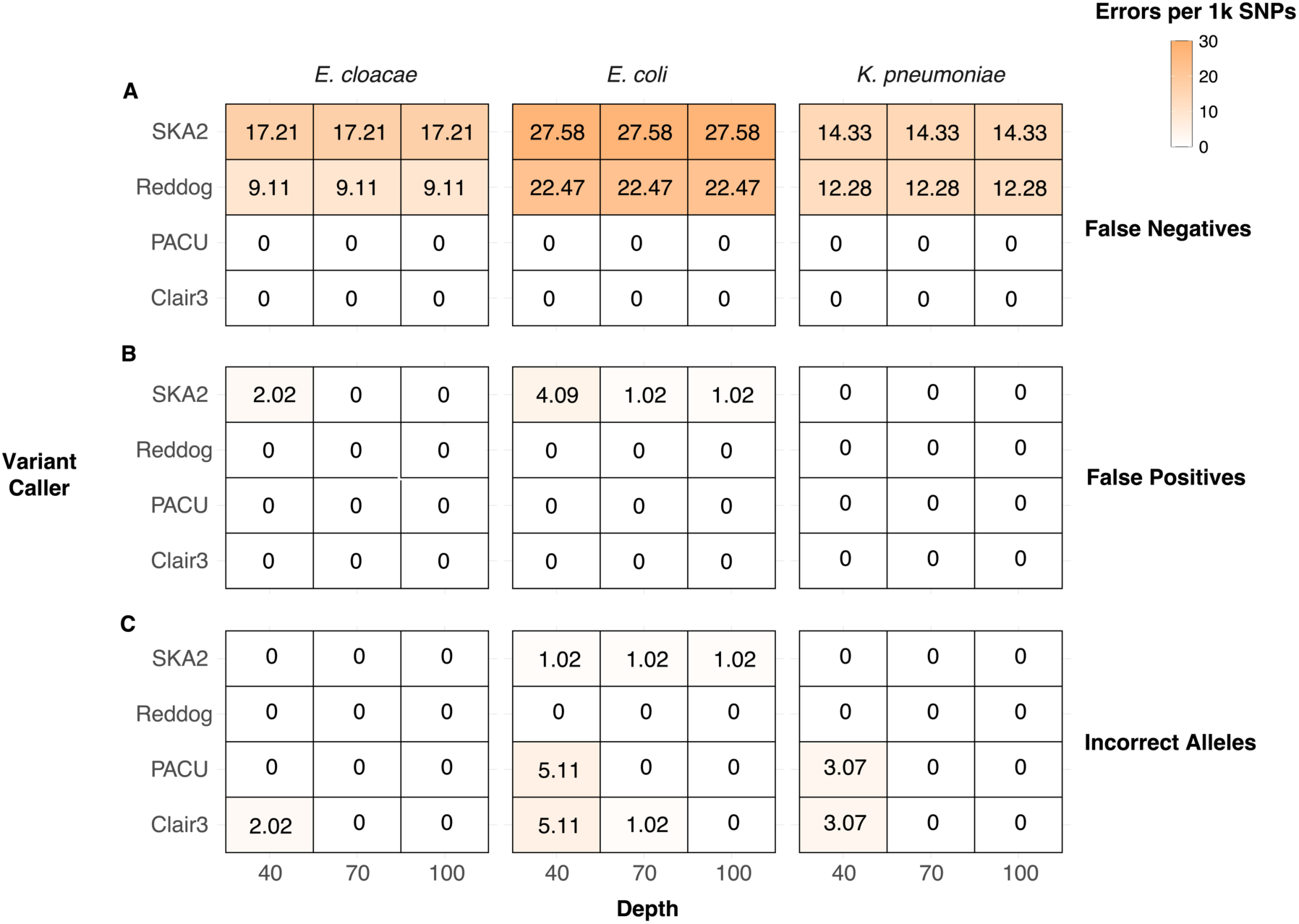
– Errors per 1000 SNPs of variant calling tools using simulated Illumina and ONT data. False negatives (**A**) refer to instances where a SNP was missed, while false positives (**B**) refer to when a SNP was called when none was present in the mutant genome. Incorrect alleles (**C**) refer to instances where a tool correctly identified the presence of a SNP at a given position in the reference genome, but the identity of the SNP was incorrect.

### Reliable SNP profiling with ONT read-based variant callers was validated with real sequencing data

We then validated our synthetic findings using real-world R10.4.1 Dorado SUP readsets, as their estimated ∼99% accuracy (3) offers the best chance for accurate variant calling. We compared pairwise SNP distributions for different variant calling tools across six *E. cloacae, E. coli,* and *K. pneumoniae* species/ST combinations (**Supp. Figures 8 – 10**, see **Methods**). While all three species showed high concordance between tools in most cases, *E. cloacae* STs showed near identical SNP distributions across all four tools (**Supp. Figure 8**). For *E. coli* ST131 **(Supp. Figure 9A**) and *K. pneumoniae* ST323 (**Supp. Figure 10B**), mapping based approaches (Reddog, Clair3, PACU) showed high concordance but SKA2 suffered from elevated SNP distances.

We then assessed sites in the reference genome that were identified as variants using different variant calling tools, while also assessing whether all strains were assigned the same allele (reference or variant) at these shared sites (**Supp. Figures 11 – 13**). We noted that while distance distributions were highly concordant across tools, the specific SNP sites and alleles calls that led to those distances were not consistent. *E. cloacae* was the most concordant species across all variant calling tools (**Supp. Figure 11**). In ST93, 84.7% (61/72) of total SNP sites in the reference genome were called in all four tools (Position Concordance in **Supp. Figure 11A**). In addition, for each site that was shared between two tools, the same allelic profile was called across all strains in nearly every case (Identity Concordance in **Supp Figure 11A**). All four variant callers were concordant in 68.8% (1979/2878) and 55.6% (15/27) of total sites for *E. coli* ST95 (**Supplementary** Figure 12A) and *K. pneumoniae* ST29 (**Supp. Figure 13A)**, respectively. *K. pneumoniae* ST323 (**Supp. Figure 13B)** showed the most discordant results due to three separate factors:

i. False positive SNP sites with SKA2 not called with any other tool (n=252 sites). This was due to a combination of random assembly errors (153/252 false positive sites) and an inability to identify heterozygous sites (99/252 false positive sites).
ii. Lowered SNP sites using Clair3 that were called using all other methods (n=20). Despite passing quality, depth and allele frequency thresholds, these sites were defined as heterozygous using Clair3 while passed as homozygous using other variant callers. Clair3 did not provide any additional information regarding these heterozygous calls, but it was likely a combination of read quality, indel misalignments and/or other extraneous factors. Clair3 offers the ability to pass heterozygous variants, but this resulted in far more false positives than false negatives and was therefore not retained in our methods. The ONT data itself was unlikely to be the issue, as PACU showed high concordance with other methods while using the same readsets.
iii. Lowered SNP sites identified using Reddog and SKA2 identified using PACU and Clair3 (n=7 sites). This was due to poor sensitivity in repetitive regions of the genome using kmer and short read methods described in the simulated read section.

Meanwhile, *E. coli* ST131 (**Supplementary** Figure 12B) showed elevated SNP sites using Clair3 (n=397 sites) and SKA2 (n=405 sites). These sites were called using all four tools but were only filtered out by Gubbins for Reddog and PACU. It is unclear why these sites made it through Gubbins for SKA2 and Clair3. However, pre-Gubbins variant calling generated over 40,000 SNPs using each tool, so it should be noted that the ∼400 SNPs that were not consistently filtered were only ∼1% of all SNPs.

### ONT mapping methods reliably identified putative transmission pairs in hospital pathogens

We then applied these results to identifying putative transmission pairs. Historically, if two Gram-negative isolates have a SNP distance of 25 or less, they are determined as putative transmission pairs (21). Of 158 pairs that met these thresholds using Reddog, ONT mapping approaches met the threshold in 155 (98.1%) and 158 (100%) of pairs for Clair3 and PACU, respectively (see **Methods**). The discordant Clair3 pairs narrowly missed the cutoff with distances of 26, 28 and 29 (**Figure 5**). Meanwhile, due to the elevated SNP sites identified using SKA2, 5/6 *K. pneumoniae* ST323 pairs did not meet the 25 SNP threshold when using SKA2.

**Figure 5.**
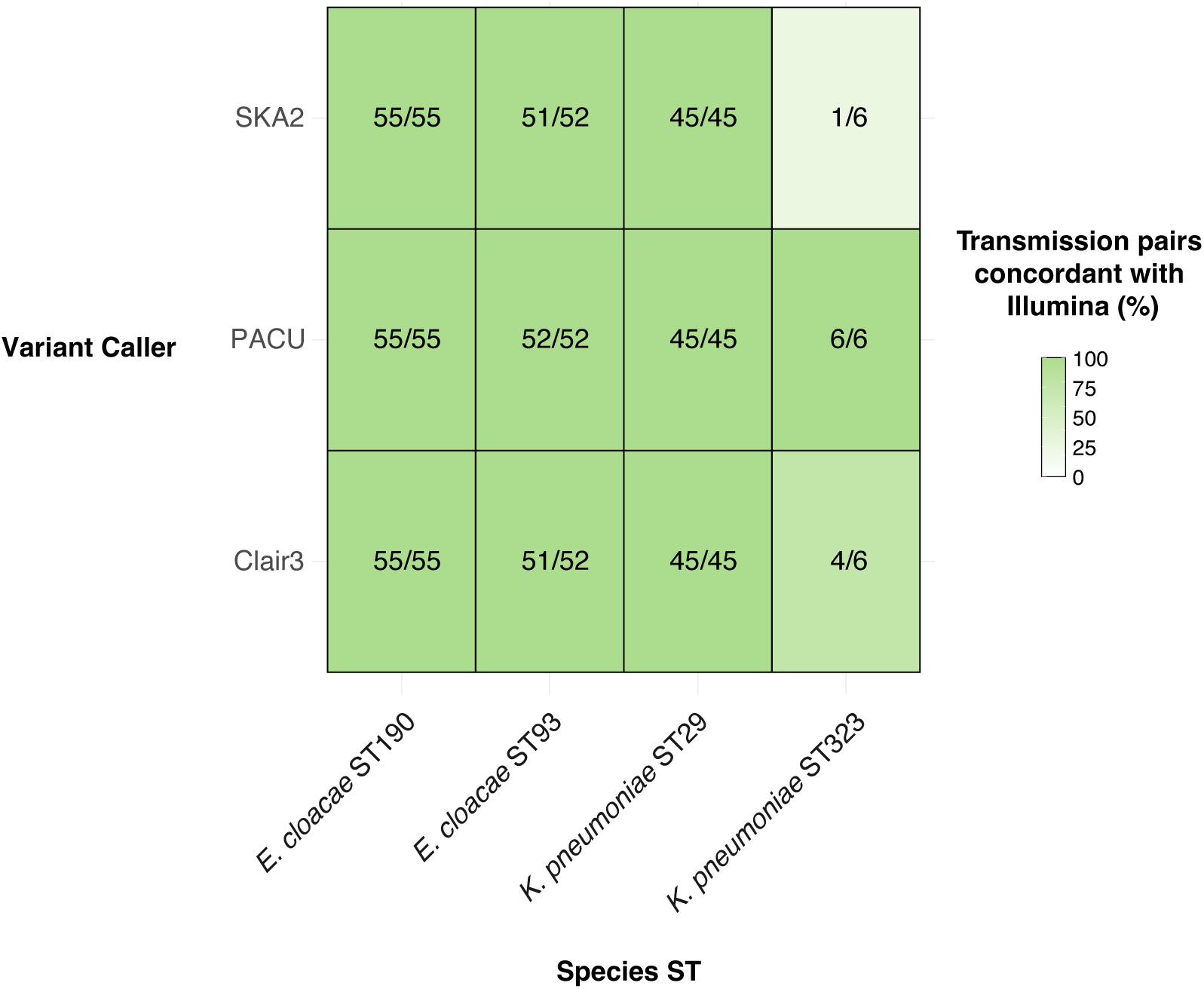
– Concordance between ONT methods and traditional Illumina methods in identifying putative transmission pairs. Included are all isolate pairs that would be assigned as putative transmission pairs using a traditional Illumina approach (Distance <= 25 SNPs). Pairs are classified as concordant with Illumina if the pairwise distance using ONT methods falls under the defined threshold for each species. No *E. coli* isolates in our dataset were putative transmission pairs so are not shown here.

We then assessed the impact of ONT sequencing on construction of SNP-based phylogenies. We compared ONT and Illumina by generating maximum likelihood phylogenies for each variant caller and compared their topologies using normalised Robinson-Fould distance and tree visualisation (see **Methods**). We noted high concordance in more diverse STs/subtrees but lower concordance in highly related STs or genetic clusters (**Supp. Table 8, Supp. Figures 14-19**). For example, *E. cloacae* ST190 showed lower topological concordance, particularly within clusters of highly related strains (**Supp. Figure 15**) while the highly divergent *E. coli* ST95 had identical topology using Reddog, Clair3 and PACU (**Supp. Figure 16**). In highly related STs, small differences in SNP sites or variant profiles between methods is enough to drastically change the topology of the tree.

### Recommendations for bacterial isolate sequencing with ONT

Due to the high error rate of legacy ONT reads, assembly has typically been a default step prior to typing analyses such as AMR, MLST and cgMLST (22). However, our work shows that that modern ONT reads perform comparably to assemblies for MLST and AMR typing, with superior AMR typing performance at depths 40x and below. Additionally, read-based tools performed better than assemblies at SNP calling due to the increased amount of quality information at each SNP site and improved performance in repetitive regions. Based on these findings, we recommend that both read and assembly-based tools are viable for AMR and MLST. At low sequencing depth we recommend read-based analyses for AMR typing, while assembly is better suited to investigating genome structure. To our knowledge there is no open-source ONT read-based cgMLST tool available, so we currently recommend assembly for cgMLST. We recommend that read-based tools such as PACU (7) or Clair3 (23) are used for variant calling over assembly-based methods.

For clinical implementation of ONT WGS, time requirements should not play a major role in deciding on assembly and analysis methods, regardless of the urgency of a result. The major factors that should be considered if time is a relevant factor (for example in the case of a hospital outbreak or severe infection) are sequencing depth and, if live basecalling is not available, basecalling model. These factors are the main determinants of how long it will take to report an actionable result; the decision of whether to assemble or not likely changes this time by no more than half an hour (**Figure 6**).

**Figure 6.**
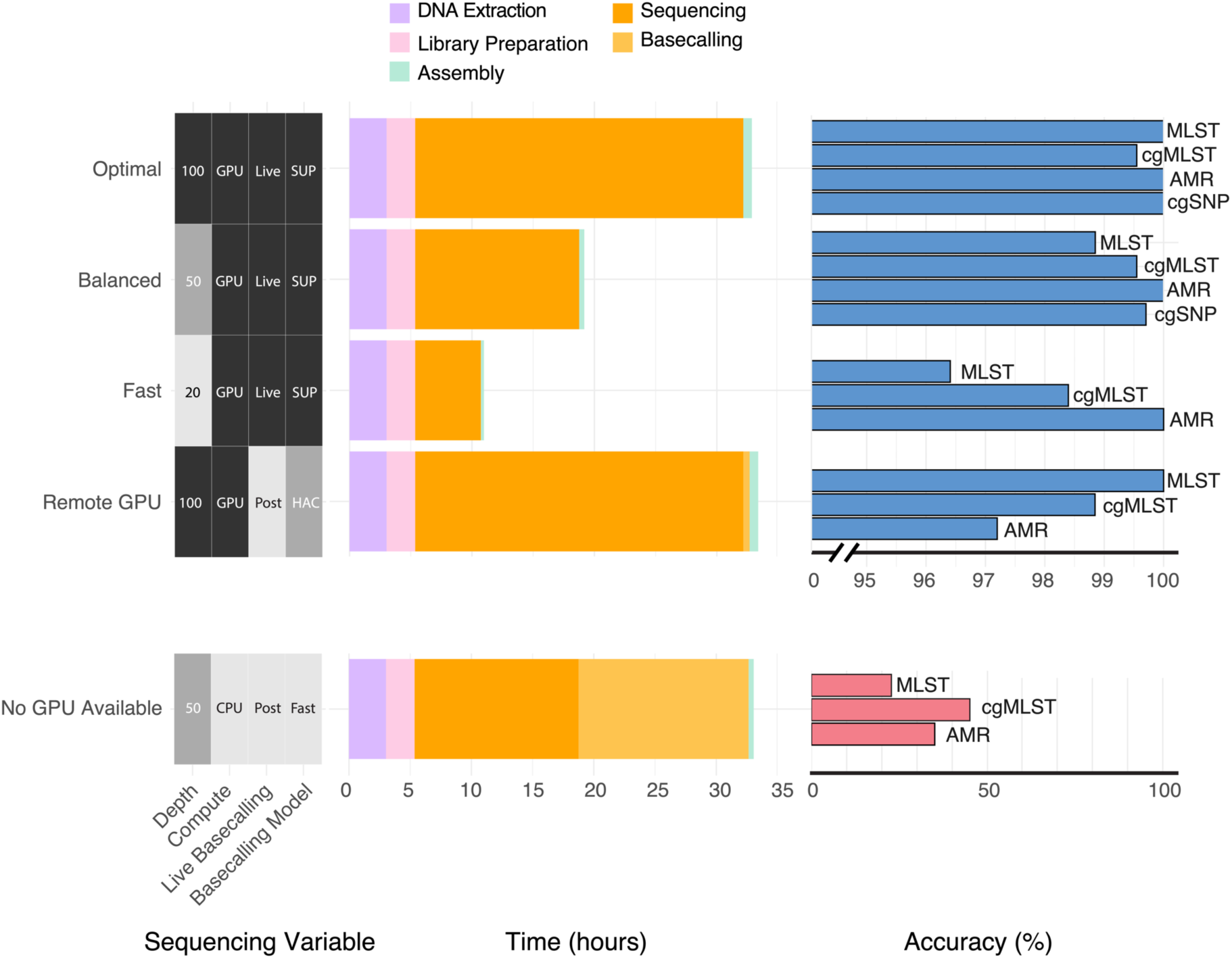
– Time requirements and expected analysis accuracy across five methods for multiplexed ONT sequencing of 20 bacterial isolates. Accuracy metrics summarise the benchmarking outcomes using R10.4.1 Dorado polished assemblies for MLST, cgMLST and AMR, except for AMR typing at 20x depth where raw reads are recommended. cgSNP accuracy was summarised from simulated R10.4.1 readsets. Accuracy metrics are coloured by GPU availability (blue=available, red=unavailable). DNA extraction and library preparation time requirements are sourced from Genfind and ONT’s protocol approximations (24, 25). Sequencing time was calculated based on all ONT runs that contributed to the benchmarking dataset. Basecalling and assembly time requirements were benchmarked on an NVIDIA A100 GPU with 60 CPUs and 40GB of RAM.

Taking these factors in consideration, we have provided five recommended combinations of sequencing depth and basecalling model. We illustrate different time and resource constraints, paired with expected turnaround times and accuracy (**Figure 6**). A more detailed breakdown of accuracy per gene and species can be found in **Supp. Figure 20**.

If a result is non-urgent and accuracy is most important, or variant calling is required, best results (“*Optimal*” in **Figure 6**) are obtained using at least 100x depth and SUP basecalling. With live basecalling, this option takes ∼33 hours when including DNA extraction, library preparation, sequencing and analysis. For faster results with high accuracy, 50x depth and SUP basecalling (“*Balanced*”) achieves 98.8-100% accuracy across all analyses with ∼17 hours required. If live basecalling is available and a rapid outcome is required, 20x depth with SUP basecalling (“*Fast*”) is likely sufficient for AMR, MLST and/or cgMLST. At 20x depth we recommend raw reads for AMR typing rather than assembly due to better benchmarking results. We are hesitant to recommend variant calling at low depths based on the known higher error rates of ONT raw reads, but did not test variant calling at these read depths. Some clinical laboratories do not have access to live basecalling, instead using a remote GPU and basecalling after sequencing. In this scenario (and assuming time to result is relevant), we recommend sequencing to 100x depth and using HAC basecalling (“*Remote GPU*”). This approach provides highly accurate typing results in ∼33 hours, with HAC basecalling requiring one fifth of SUP basecalling time based on our benchmarking (**Supp. Table 9**). Finally, if there is no GPU access (“*No GPU*”), we do not recommend attempting typing analyses due to poor accuracy of the Fast basecalling (the only viable model to use without a GPU).

## Discussion

In this study, we demonstrate that modern R10.4.1 ONT data is now a viable choice for characterisation of bacterial pathogens in clinical settings. We noted very high concordance to genomic gold standards and comparable or better performance than Illumina. R10.4.1 sequences basecalled with the SUP model enabled perfect MLST and AMR calls across 85 *Enterobacterales* isolates. SUP datasets had a median cgMLST locus distance of just 11 genes to the gold standard genomes of KpSC and *E. coli* isolates. ONT read-mapping variant callers outperformed traditional Illumina methods and kmer-based methods using simulated read data due to improved performance in repetitive regions of the genome. ONT mapping approaches showed high concordance with Illumina approaches using real sequence data, generating similar distance distributions, SNP sites and identification of putative transmission pairs. Phylogenies constructed with ONT mapping-based methods resembled those generated with Illumina for diverse STs, but often showed several topological changes in STs with smaller genetic distances due to the increased impacts of any singular discordant SNP call. Our findings have allowed us to provide specific recommendations for clinical ONT sequencing regarding depth, basecalling model and analysis methods depending on the goals and resources available to the user.

To our knowledge, a single prior study (6) has tested MLST accuracy of modern ONT data. This study also noted perfect concordance between Illumina and ONT for all assemblies containing a full MLST profile. 5% of ONT assemblies in this study were missing one or more MLST locus, similar to the 3.7% (10/268) isolates removed from our benchmarking dataset due to unresolved MLST profiles in gold standards. In this situation, read-based MLST profiling or an alternative assembly algorithm (e.g. Hybracter (26) or Autocycler (27)) should be used. We found cgMLST accuracy of our ONT-only datasets (median of 11 cgMLST loci incorrect per isolate) to be comparable to other studies that followed a similar approach of Flye assembly followed by Medaka polishing (8, 10–12). Some studies have sought to further optimise this accuracy through using methods such as duplex ONT sequencing (6), specific cgMLST optimisation tools (12) and bacteria-specific Medaka models (8). While these optimisations were outside of the scope of this study, they found that cgMLST performance increased to near-perfect (0-2 gene distance to gold standard), suggesting that our increased threshold for defining cgMLST clusters (**Figure 3**) may not be necessary with future advancements.

The excellent performance of ONT-only assemblies in identifying AMR allelic variants shown here is comparable to recent benchmarking performed in twelve Gram-negative isolates (14). However, the imperfect retention of AMR genes in Flye assemblies at varying depths (**Supp. Figure 7**) is something that should be monitored going forward. While Landman et al. (6) did test for AMR gene retention and found very few issues, the isolates they used had fewer AMR genes (six genes across all isolates, compared to 17 genes in our dataset). Going forward, multi-input assembly algorithms such as Autocycler (27) and Hybracter (26), or a combination of read and assembly-based approaches, may offer a more robust method for identifying and typing AMR allelic variants whilst also investigating their genomic context.

The poor sensitivity of variant calling in repetitive regions using Reddog and SKA2 shown here was in line with recent research by Hall et al. (5), who demonstrated that Illumina approaches could not reliably resolve repetitive regions longer than its short read lengths. A similar limitation is discussed in the SKA2 paper (28), where they acknowledge that repeat regions longer than the max kmer length can lead to false negatives. False positive SNPs observed in this study were dominated by SKA2, due to an inability to identify heterozygous sites or errors introduced during the assembly process. In STs where this occurred, such as *E. coli* ST131 **(Supp. Figure 12B**) and *K. pneumoniae* ST323 **(Supp. Figure 13B**), the number of these SKA2-only sites was roughly equal to the consensus error rate (∼0.5 errors per 100kbp) of modern ONT data (29). These errors point to limitations of assembly rather than SKA2 itself. Assemblers do not provide quality information that would allow for identification of heterozygous sites and filtering of sites with low quality scores, sequencing depth or similar. In addition, SKA2 was designed for Illumina data with a higher consensus accuracy and may not be suited to more error-prone ONT data. For these reasons, read mapping is a more reliable method for variant calling from ONT data.

This study had three potential limitations surrounding the dataset used. The first is that ONT and Illumina sequencing were performed on different extracts, potentially allowing the bacterial strain to mutate between extractions. This did not appear to have a major impact on any of the benchmarking, as we found ONT accuracy to be comparable to other benchmarking studies in most cases. In instances where ONT datasets performed worse than previously observed or expected, these discrepancies could be explained by a combination of lack of assembly optimisation (cgMLST), inherent limitations of assembly for variant calling (SKA2) or Clair3-specific inconsistencies not observed in the other ONT read-based variant caller (PACU). The second limitation surrounds the basecalling and assembly algorithms used – while Flye was the best performing fully automated assembly algorithm when we began this analysis (30), Hybracter (26) and Autocycler (27) have since been released which combine the strengths of multiple assembly algorithms at once in a fully automated fashion. Autocycler or Hybracter assemblies may lead to lowered rates of presumable false positives with SKA2 and improved cgMLST performance due to a lower error rate in assembly overall. Future benchmarking should look to include these algorithms as well as the new bacteria-specific Dorado and Medaka models to further optimise accuracy. Finally, this study focused on common Gram-negative pathogens. Future studies should validate benchmarking in Gram-positive and less common bacteria taxa, as ONT benchmarking algorithms may not have been trained to perform as well in these cases.

The benchmarking shown in this study highlights the extremely high accuracy of modern R10.4.1 ONT data and its improvement over legacy ONT chemistries and software. While assembly has historically been a default step prior to MLST or AMR typing, we found that read-based tools may be better equipped to avoid omissions of MLST loci or plasmid-associated AMR genes and provide superior variant calling performance due to detection of heterozygous loci and avoiding assembly errors. ONT long reads showed improved performance over Illumina in capturing repetitive regions of the genome and had comparable performance to Illumina outside of repetitive regions, suggesting that they may be the best available method for characterising outbreaks in hospital settings. When combining the improved accuracy of ONT sequencing with its existing cost and analysis benefits, it appears poised to become the definitive method for characterising bacterial isolates in clinical settings going forward.

## Methods

### Bacterial isolate collection and sequencing

We chose 268 KpSC, ECC and *E. coli* strains from our institutional biorepository with existing Illumina sequencing data and performed additional ONT sequencing (**Table 1, Supp. Table 1**). All DNA extractions were done using GenFind kits according to manufacturer guidelines (31). ONT sequencing was performed on new DNA extracts since it was typically done several years after the original Illumina sequencing. Illumina sequencing was performed on the HiSeq 2000 and Novaseq 6000 platforms, while ONT sequencing was performed on R9.4.1 and R10.4.1 flowcells using the ligation (LSK-109 for R9.4.1) and native (NBD-196 for R10.4.1) barcoding kits.

### Basecalling and assembly

Raw ONT fast5 files were processed in several steps including basecalling, read subsetting, assembly and characterisation using commonly used tools (**Figure 1**). All fast5 files were basecalled using ONT’s legacy basecaller Guppy v6.2.1 and R10.4.1 sequences were also basecalled using the newer software Dorado v0.5.1 (32) software. Separate fastq files were generated with all three basecalling models (SUP, HAC and Fast). R9.4.1 basecalling was performed on an NVIDIA T4 graphics processing unit (GPU) with 16 CPUs and 64GB of RAM. R10.4.1 basecalling was performed on an NVIDIA A100 with 60 CPUs and 40GB of RAM or the NVIDIA V100 GPU present in ONT’s gridION device.

To ensure we were using high quality data, we first filtered out any isolates with sequencing depth <40x (n=48 R9.4.1 isolates, n=6 for R10.4.1). We then ran proch-n50 v1.5.8 (33) to capture read length and NanoStat v1.6.0 (34) to capture median read quality; both with default settings. Read length was high (median 22,469 bp, IQR 17,219 – 28,067), and read quality was in line with typical scores relative to sequencing chemistry and basecalling algorithms (**Supp. Figure 1, Supp. Table 1**) (3, 14). As expected, R10.4.1 reads (median quality score of 17.2 – 20.1 using Dorado SUP) were of higher quality than R9.4.1 reads (median quality score of 11.2 – 15.8 using SUP) and quality increased from Fast to HAC to SUP models (**Supp. Figure 1**).

We then used R9.4.1 Guppy and R10.4.1 Dorado SUP basecalled fastq files to generate hybrid gold standard assemblies using a custom-built nextflow pipeline (35) based on the assembly method recommendations in the Trycycler (36) and Polypolish (37) studies. The pipeline filters reads using Filtlong v0.2.1 (38) with the parameters –-min_length 1000 and –-keep_percent 95, then assembles using Flye v2.9 (39) with the –-nano-hq tag. It then performs long read polishing using Medaka v1.7 (40) with a user specified model (-m) depending on the input dataset and short read polishes using Polypolish v0.5.0 (37) and Polca v3.4.2 (41) with default parameters if short read data is available. We used contig number and MLST calls as quality control (QC) metrics for filtering out poor quality gold standard assemblies. Any isolates with more than 20 contigs in the gold standard assembly (n=0 for R9.4.1, n=2 for R10.4.1) and/or missing/incomplete sequences in any MLST loci for gold standard assemblies (n=2 for R9.4.1, n=7 for R10.4.1) were excluded. We also excluded three R9.4.1 isolates due to obvious isolate mismatches between ONT and Illumina data resulting from mislabelling or errors in the sequencing process. The final benchmarking dataset was comprised of 199 isolates (**Table 1, Supp. Table 1**).

To generate subset assemblies, we randomised the order of reads in full fastq files then took increasing proportions of the full readset to reach depth levels from 10x – 100x using the subseq command of Seqtk v1.4 (42). The order of reads was consistent between depth levels to ensure that each increasing depth level contained the same reads as the last, plus an additional set to reach the next depth level. These subsetted readsets were then fed into the nextflow pipeline (35) for assembly. Reads generated from all three basecalling models (Fast, HAC and SUP) were processed in this way. Since Medaka does not have a model designed for Dorado Fast basecalling, we used the HAC model to perform long read polishing of Fast assemblies.

### Typing analyses

For gold standard and subset assembly MLST calls we used mlst v2.23.0 (43) with the relevant –s flag dependent on the organism being analysed. We used bart v0.1.2 (15) with the –r ont flag and appropriate –s species flag for read-based MLST.

For cgMLST We used chewBBACA v3.3.10 (19) using the *E. coli* and KpSC databases from cgmlst.org (44). This involved downloading the species database fasta file, running the PrepExternalScheme command with default settings, the AlleleCall command on gold standard assemblies while allowing inferred alleles to be added to the database (default settings), then running AlleleCall on the subset assemblies without adding inferred alleles to the database (--no-inferred) to ensure that each group of subset assemblies was run on the same database. To assign strains to clusters based on cgMLST typing, We used cgMLST-dists v0.4.0 (45) with default settings to generate distances matrices and then assigned strains to clusters based on average distance thresholds using hclust (46) in R. To visualise these clusters we used igraph (47) and ggplot2 (48) in R.

For AMR calling from hybrid assemblies, we ran AMRFinderPlus v3.12.8 (49) using default settings and the NCBI curated database. For subset assemblies, we used an identity threshold of 70% to avoid false negatives in low depth assemblies that were expected to be of poor quality. When comparing subset performance to the gold standard for determining AMR allelic variants, we only used strains for which an allelic variant was called using AMRFinderPlus on the gold standard assembly. We used bart v0.1.2 (15) with the –r ont flag and additional –amr flag for read-based AMR calling.

To compare the MLST, cgMLST and AMR profiles of ONT-only data inputs to genomic gold standards of the same isolate, we removed confidence symbols from the tool outputs and summarised the number of correct alleles using string-matching in R. For read-based analyses, bart often reports several possible allelic combinations in order of decreasing confidence. For MLST, we took the top match. For AMR, we took the top n hits per gene by alignment depth, where n represents the number of allelic variants of that gene present in the gold standard assembly. In most cases n=1, however some gold standards contained *bla*_CTX-M-14_ and *bla*_CTX-M-15_, for example. For these isolates we took the top two hits by depth from the bart output. For cgMLST analyses, we removed loci with no allele call in 5 or more gold standard assemblies (n=176 for KpSC, n=93 for *E. coli*) as well as isolates with ten or more loci with no allele call (n=2 for KpSC and *E. coli*).

To calculate AMR gene retention rate in subset assemblies, we first extracted all AMR genes identified by AMRFinderPlus in the gold standard assemblies from the AMRFinderPlus database using Seqtk v1.4 (42). We then aligned all subset assemblies to those genes using Minimap2 v2.26 (18) using the –-secondary=no flag and reported whether they generated an alignment or not. All benchmarking results were parsed and plotted in R using the dplyr (50), reshape2 (51) and ggplot2 (48) packages.

### Simulated data preparation for variant calling

Determining the ground truth in highly sensitive analyses such as variant calling can be difficult due to imperfections in any assembly method (36). To test variant callers with an established ground truth, we began by benchmarking performance with simulated reads. This included selecting a random *E. cloacae, E. coli,* and *K. pneumoniae* hybrid assembly and introducing 1000 random SNP sites into its chromosome using a custom Python script. The commands and scripts used to perform and analyse variant calling are publicly available at https://github.com/HughCottingham/ONT_benchmarking_paper. We generated ten mutant assemblies per reference, where each mutant assembly had SNPs in 0-1000 of these sites.

We then simulated ONT and Illumina raw reads for each of the mutant genomes. For ONT, we simulated reads at 40x, 70x and 100x depth using NanoSim v2.0.0 (52). This involved training the simulation model using the read_analysis.py genome command with default parameters and the hybrid reference genome with its corresponding R10.4.1 Dorado SUP reads as inputs. Next we ran NanoSim’s simulator.py genome command with the following parameters: –rf mutant_genome.fasta, –c training_file –n genome_size/100 (or 70 or 40 depending on fold coverage) –-fastq –med 10000 –sd 0.3. We generated 100x Illumina HiSeq reads using ART v2016.06.05 (53) with the following parameters: –ss HS20 –i mutant_genome.fasta –p –m 300 –l 100 –s 50 –f 100. We assembled all simulated ONT readsets as described earlier using our nextflow pipeline (35).

### Variant Calling

Once we had both Illumina and ONT R10.4.1 Dorado SUP reads (simulated and real), we used the chromosome of one gold standard genome per ST as a reference and called SNPs using four different variant callers (**Supp. Table 10**). We used Reddog (54) to call variants from Illumina reads, which has been extensively used across several previous studies (55, 56). Reddog aligns reads to a reference genome using Bowtie 2 v2.4.1 (57), calls SNPs using Samtools v1.9 (58) and reports variants with quality score >=30. For ONT assemblies, we chose to assess performance using the split-kmer tool SKA2 due to its simplicity for future clinical utility combined with high reported accuracy with Illumina assemblies (28). We used the ska build and ska map commands from SKA2 v0.3.7 (28) with default settings as described in their documentation. For ONT reads, we used PACU v0.0.6 (7), which was the only pipeline to our knowledge specifically built for cgSNP typing from R10.4.1 ONT reads. PACU calls SNPs in the reference genome using BCFtools v1.17 and filters for quality score and low depth regions in a similar fashion to Reddog and other established cgSNP tools. Additionally, we also assessed performance of the Clair3 v1.0.10 (23) variant caller when combined with our own manual filtering, as this has recently found to be most accurate variant caller for R10.4.1 reads (5). We initially set allele frequency (AF) to 0.9 and the quality threshold to 30 for PACU and Clair3 to mirror Reddog. However, we found these thresholds to be too stringent due to the differences in the sequencing platform and variant calling process. Instead, we relaxed the quality and AF thresholds to better suit each tool and improve concordance (**Supp. Table 10**). In addition to these adjustments, we ran Clair3 using the r1041_e82_400bps_sup_v430 model found on Rerio (59) and included the parameters –-haploid_precise to filter out heterozygous calls and –-no_phasing_for_fa to skip phasing, which is not relevant in haploid organisms. For benchmarking variant calling performance, we chose to focus on SNPs due to their clinical utility in calculating pairwise distance matrices rather than other metrics such as indels and large structural variants.

Each of these tools outputs a variant call format (VCF) file showing variant sites in the reference genome. Using a custom Python script, we generated whole genome alignment files where each sequence was a copy of the reference genome with SNPs introduced according to those identified in each of the strains. We then fed this whole genome alignment file into Gubbins v3.4 (60) with default parameters. Finally, we converted the VCF output of Gubbins into final allele matrices using another custom Python script. The commands and scripts used for these processes can be found at: https://github.com/HughCottingham/ONT_benchmarking_paper.

To compare pairwise distances between methods, we generated fasta alignment files from the allele matrices using a custom Python script and fed those files into snp-dists v0.8.2 (61) with default settings. We then plotted these distances in R using dplyr (50) and ggplot2 (48). We summarised concordance of SNP sites using upsetR (62) with the allele matrices as input and summarised identity concordance using dplyr and ggplot2. To determine whether SNP sites were located in repeat regions of the reference genome, we followed the approach outlined by Hall et al (5) built on using mummer to align the genome to itself and output alignments with identity 60% or greater. To determine concordance in identifying putative transmission pairs, we used a 25 SNP threshold (21) for Gram-negative bacteria. We did not have any *E. coli* strains within these thresholds, so they were not part of this analysis. To generate phylogenies, we ran RAxML-NG v1.2.1 (63) on the core genome alignment fasta files with the following parameters: –-all –-model LG+G8+F –-tree pars10 –-bs-trees 200. To calculate the similarity of tree topology between variant callers we used normalised Robinson-Fould distance, which describes homology from 0 (identical) to 1 (no similarity) by calculating the number of splits in topology / total splits across both trees. We used ETE3 v3.1.3 (64) to perform midpoint rooting and distance calculations.

## Data availability

All data used in this study are freely available via Figshare (https://doi.org/10.26180/c.7942409.v1). Additionally, Illumina reads and the highest quality ONT reads have been deposited in the SRA or ENA, with accessions listed in **Supplementary Table 1.**

## Acknowledgements

This research was supported by Monash eResearch capabilities, including M3.

This research was also supported by use of the Nectar Research Cloud, a collaborative Australian research platform supported by the NCRIS-funded Australian Research Data Commons (ARDC).

