## Supplemental Figures for "Nanopore sequencing enables highly accurate genotyping and identification of resistance determinants in key nosocomial pathogens"

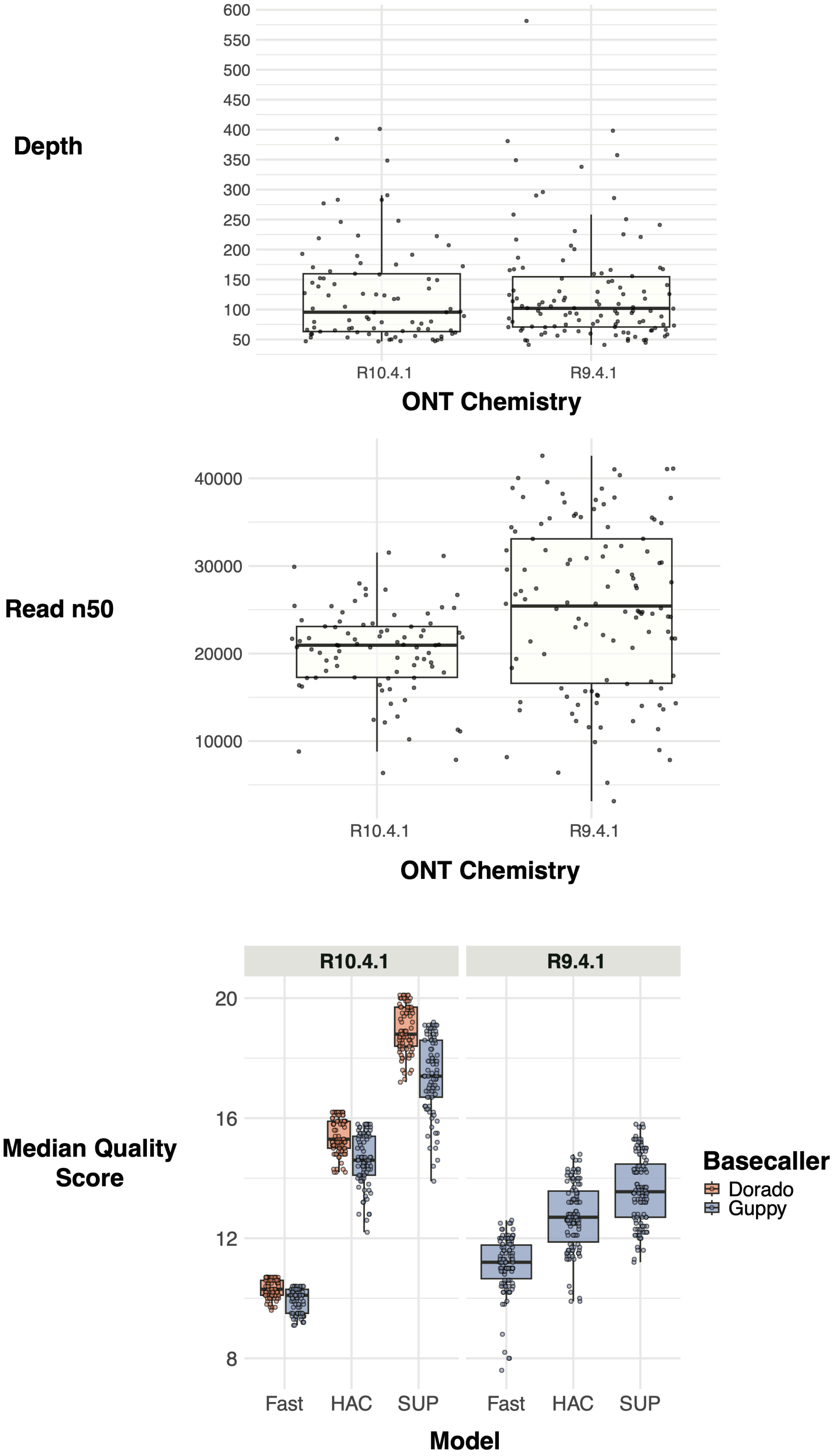


**Supplementary Figure 1 – Summary of read depth, length and quality in the final benchmarking dataset.**

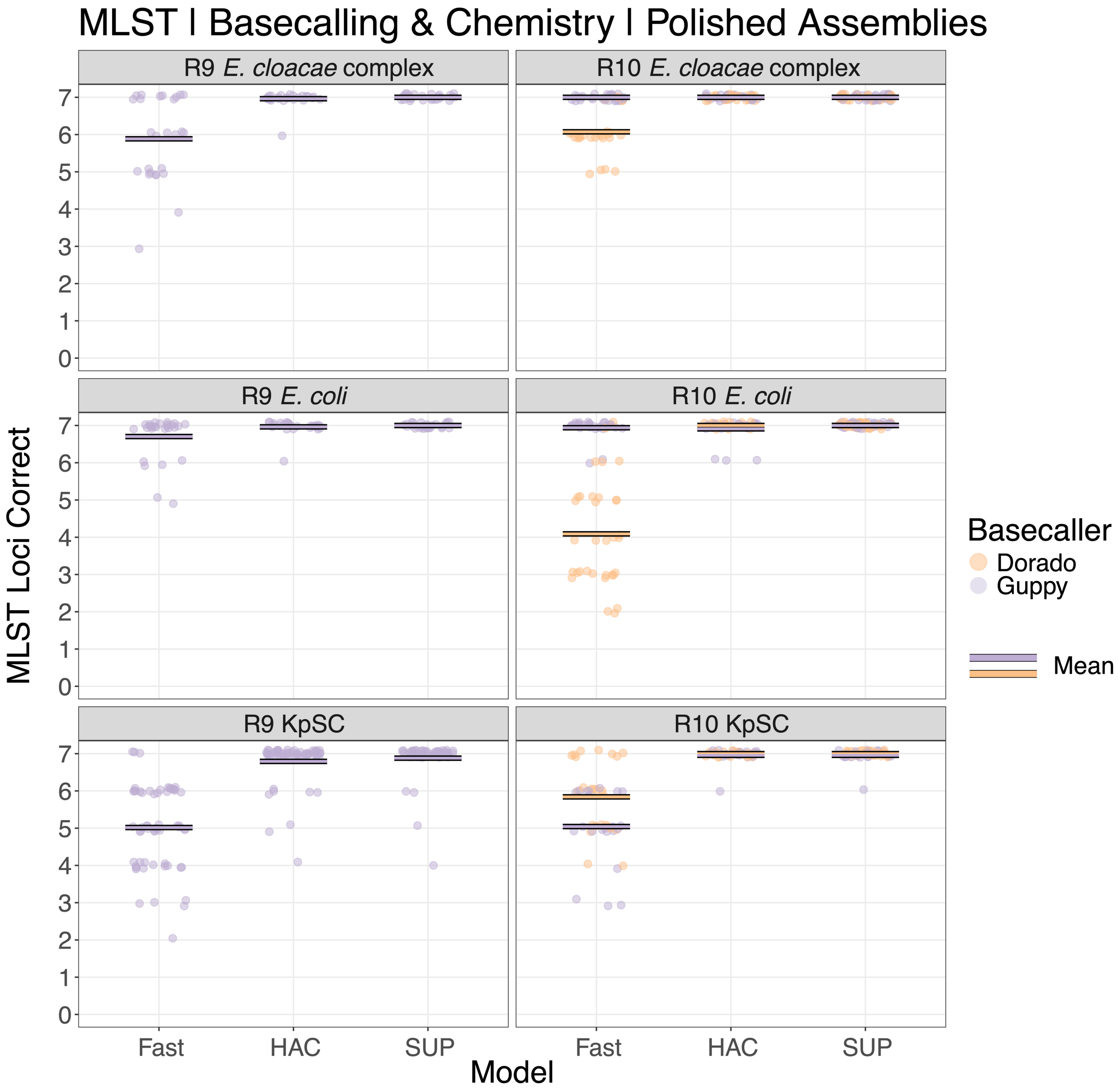


**Supplementary Figure 2 – Accuracy of MLST genotyping using polished ONT-only assemblies generated from the maximal depth available for each isolate (max 100x depth).** Accuracy is broken down according to legacy R9.4.1 and modern R10.4.1 chemistry as well as basecalling model and software.

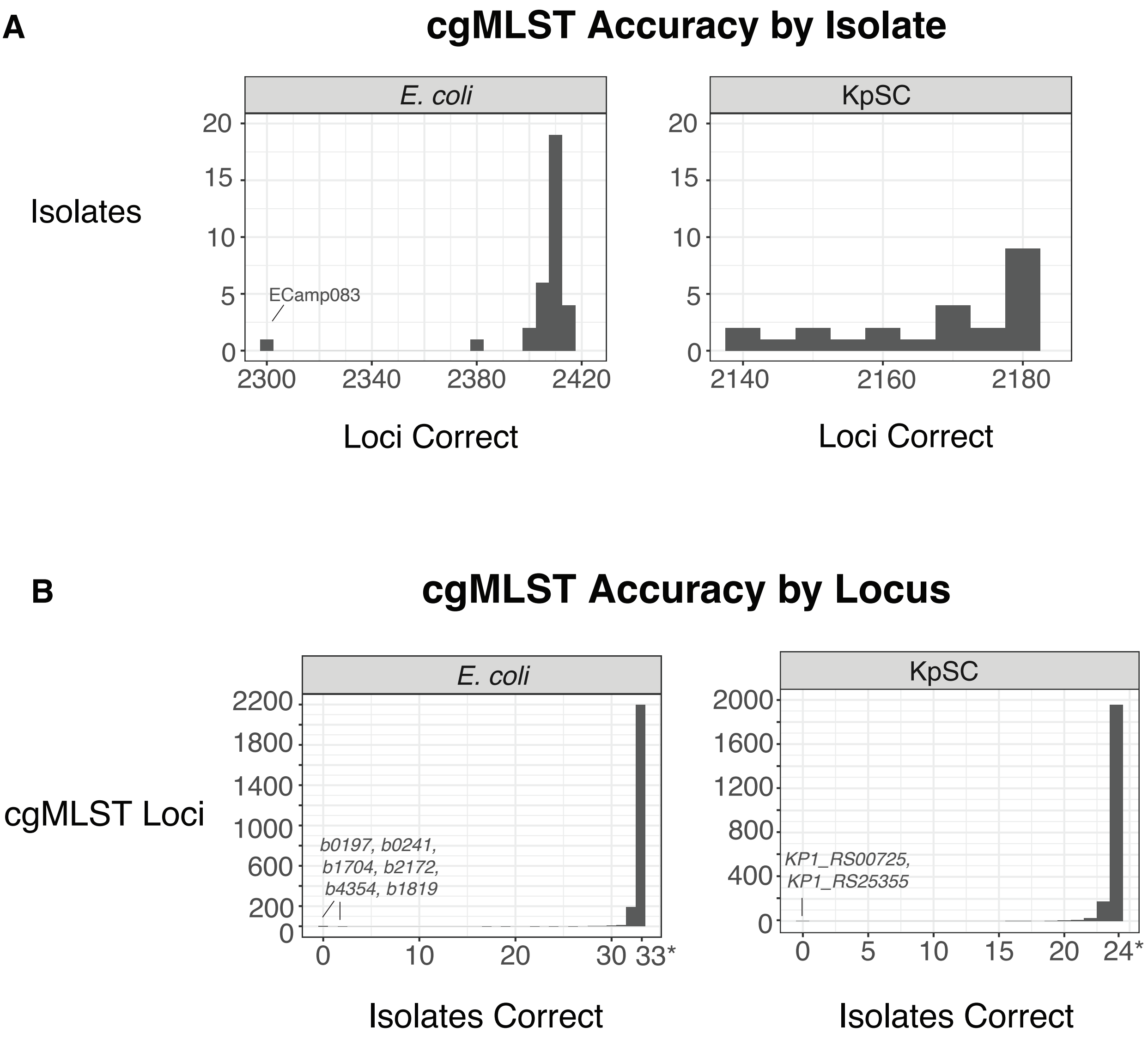


**Supplementary Figure 3 – Accuracy of *E. coli* and KpSC cgMLST genotyping using the maximum depth available (100x or below) for each strain, basecalled using Dorado SUP model, and assembled with Medaka polishing**. Isolates and loci with poor allele calling accuracy are labelled. **A)** Number of correct cgMLST loci by isolate **B)** Number of isolates with a correct call by cgMLST locus. *Maximum number of isolates included in benchmarking.

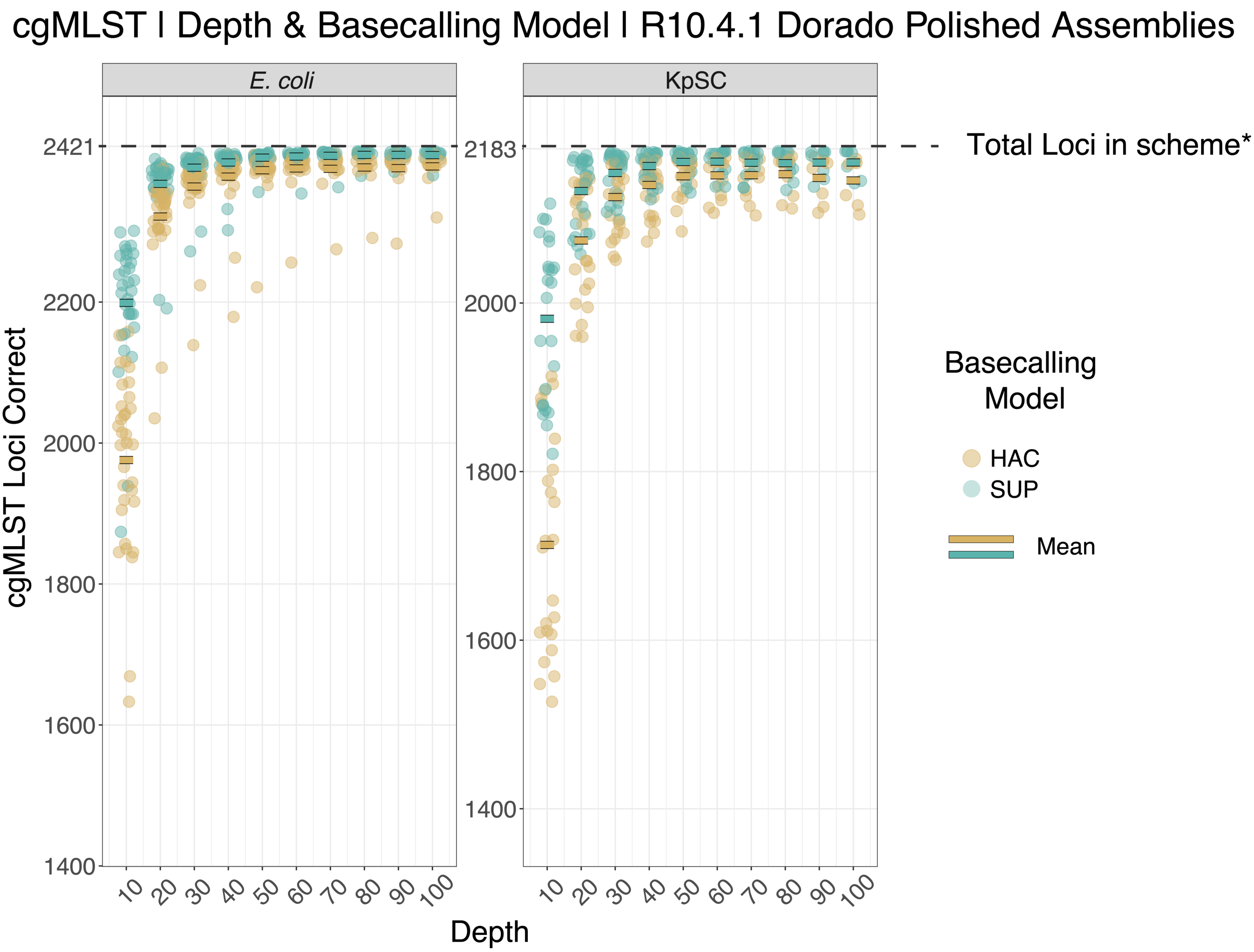


**Supplementary Figure 4 - Accuracy of *E. coli* and KpSC cgMLST genotyping using HAC and SUP Medaka-polished assemblies from 10x – 100x depth (when available).** *After filtering loci poorly defined gold standard genomes.

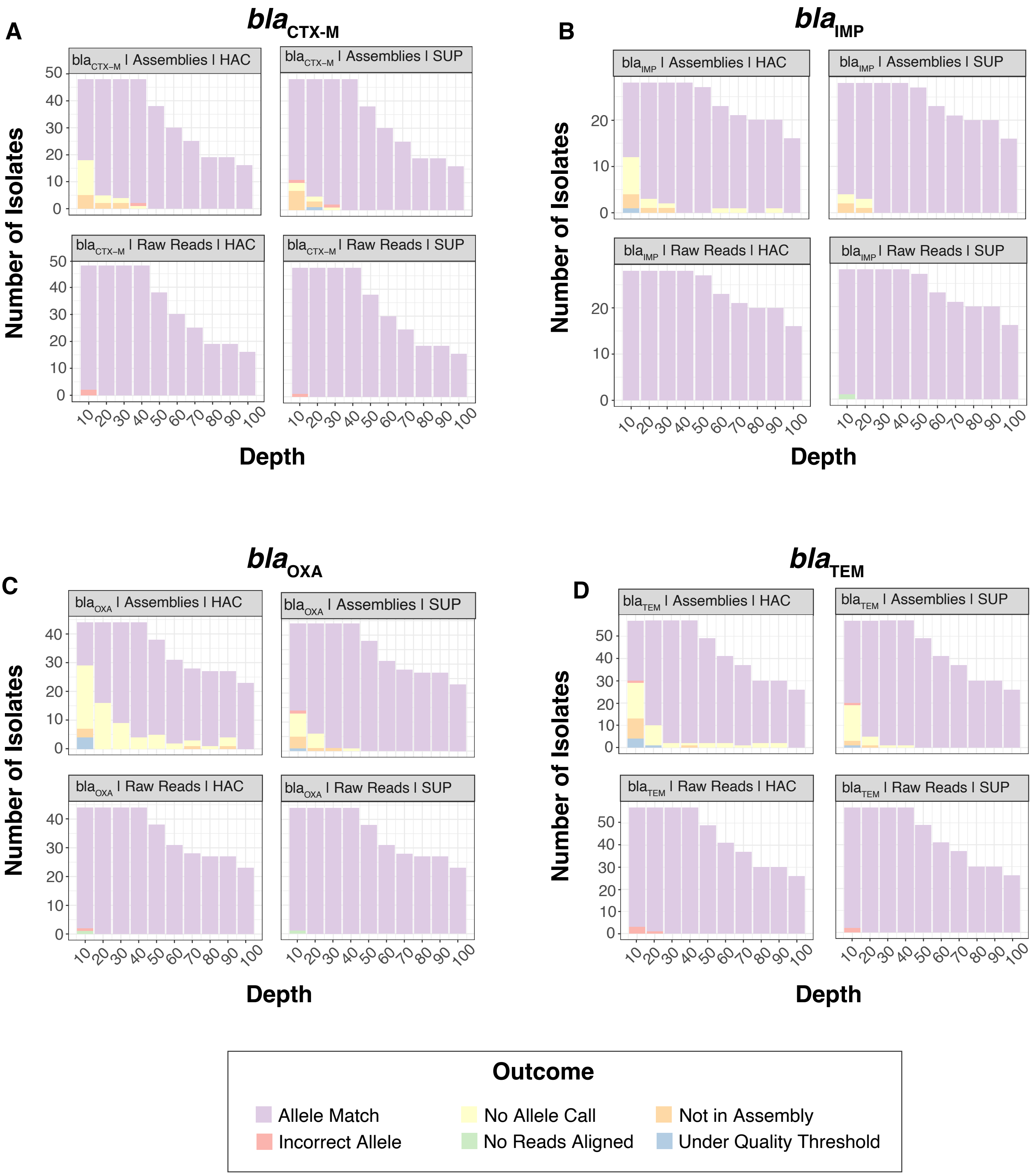


**Supplementary Figure 5 - Accuracy of AMR allelic variants using modern ONT-only data at varying sequence depths**. **A)** Accuracy across bla$_{CTX\text{-}M}$ alleles. **B)** Accuracy across bla$_{IMP}$ alleles. **C)** Accuracy across bla$_{OXA}$ alleles. **D)** Accuracy across bla$_{TEM}$ alleles.

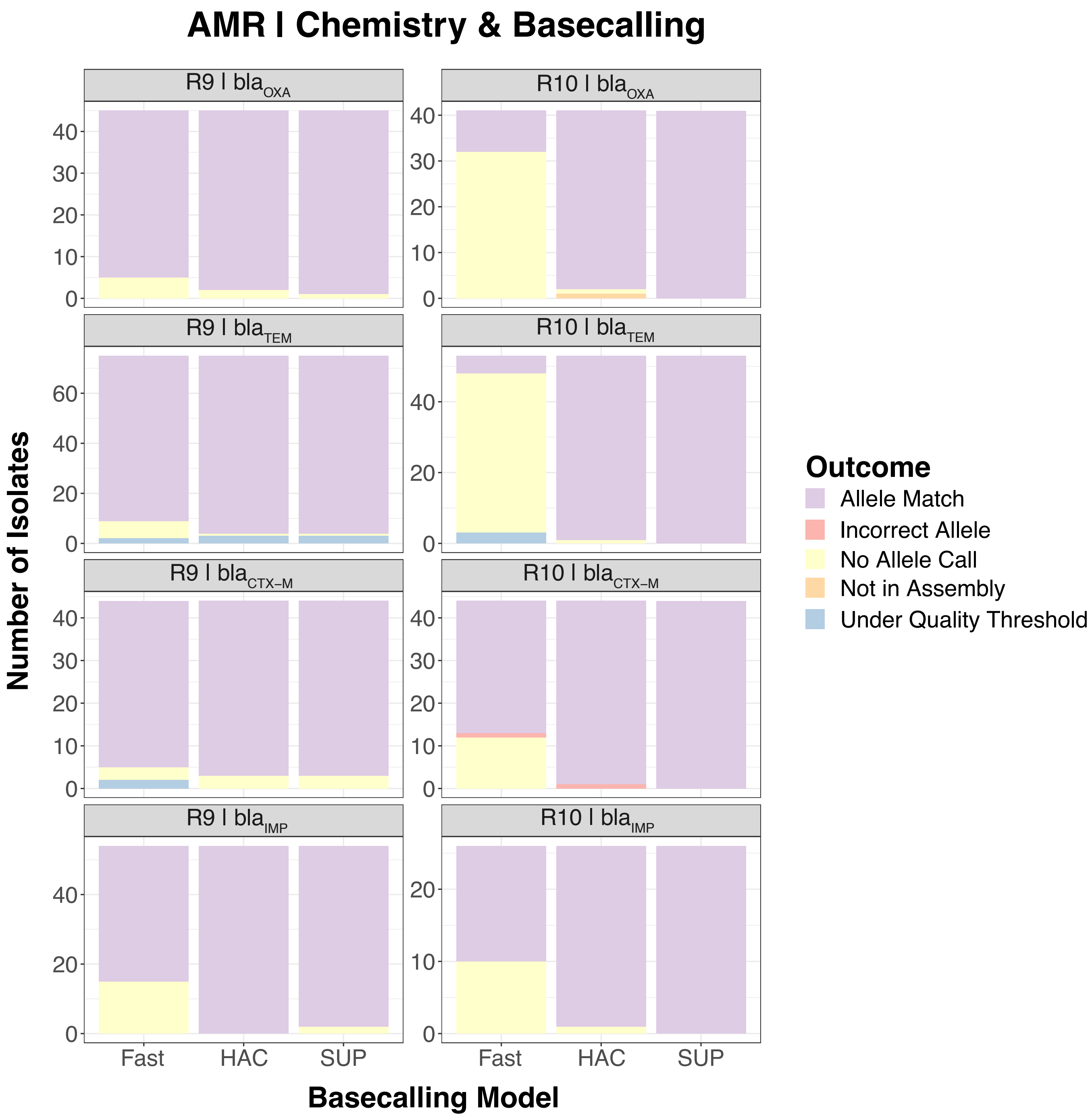


**Supplementary Figure 6 - Accuracy of AMR allelic variants using polished ONT-only assemblies generated from the maximal depth available for each isolate (100x and below).** Accuracy is broken down according to legacy R9.4.1 and modern R10.4.1 chemistry as well as basecalling model and software.

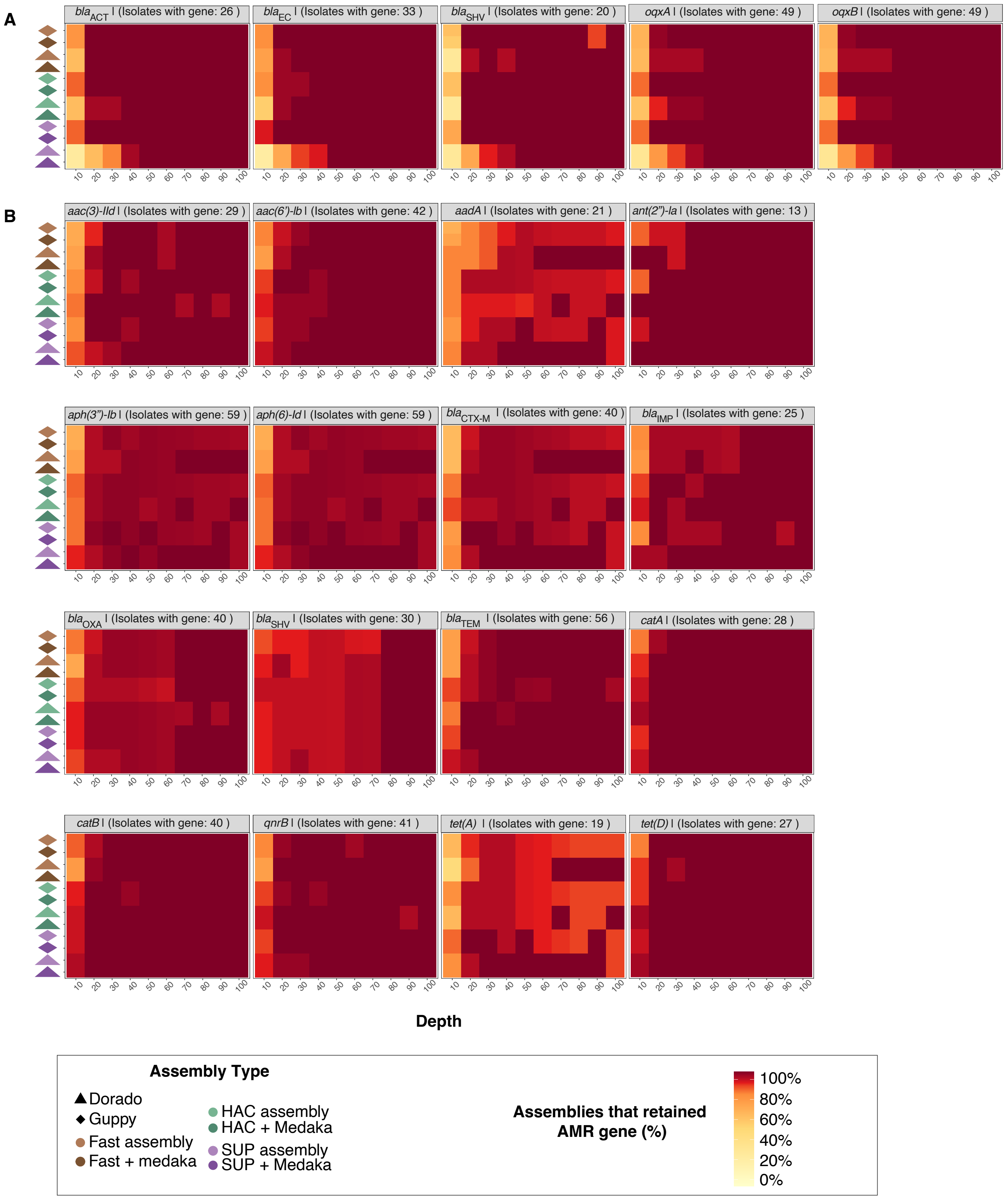


**Supplementary Figure 7 - Retention of AMR genes in A) chromosomes and B) plasmids following varying assembly methods with R10.4.1 ONT-only data types.** Shown are genes present in the gold standard genomes of at least ten isolates.

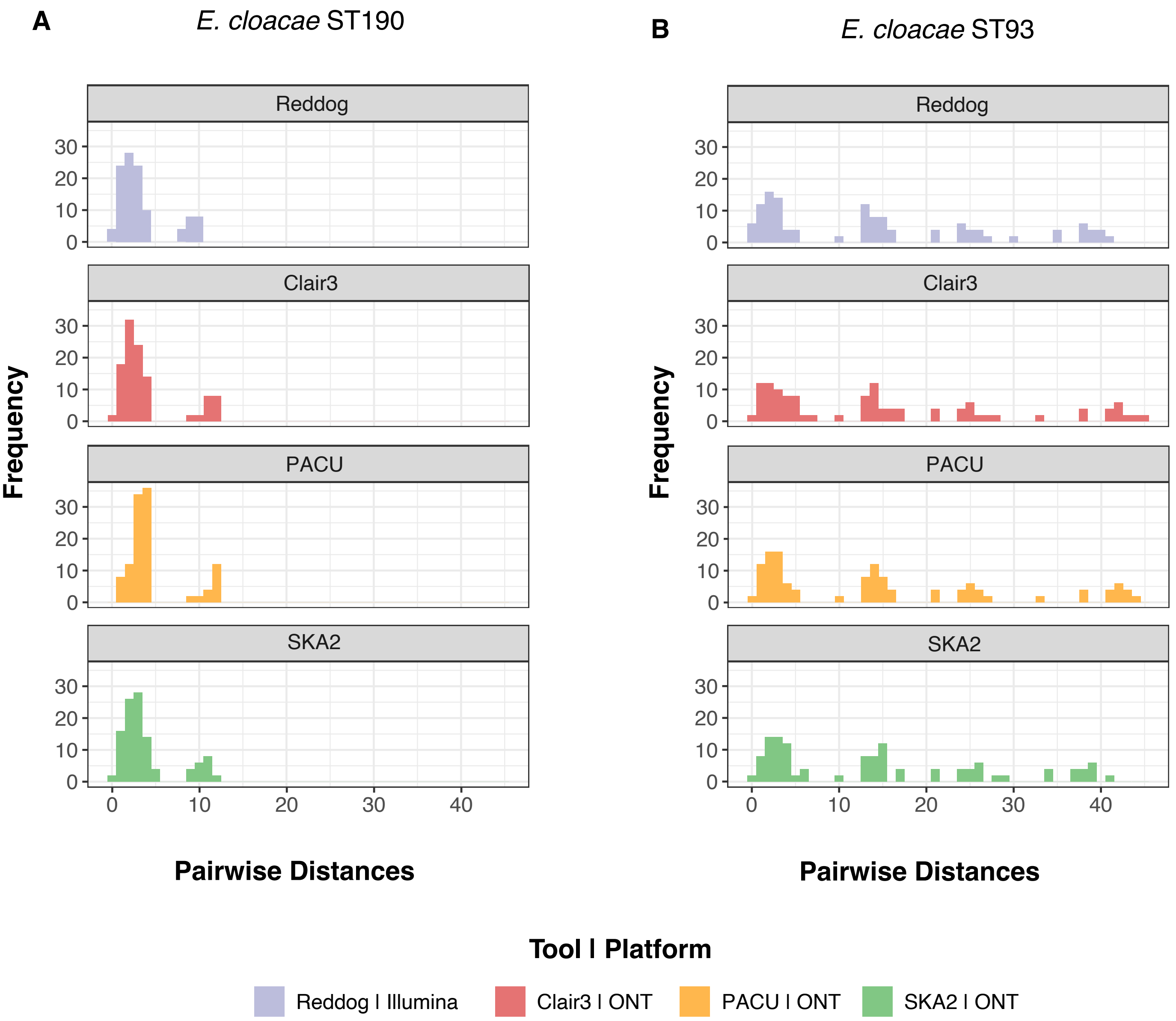


**Supplementary Figure 8 - Pairwise distance distribution of *E. cloacae* isolates after calling core genome SNPs using varying variant calling tools.** Distributions are coloured by the tool used. **A)** *E. cloacae* ST190 isolates. **B)** *E. cloacae* ST93 isolates.

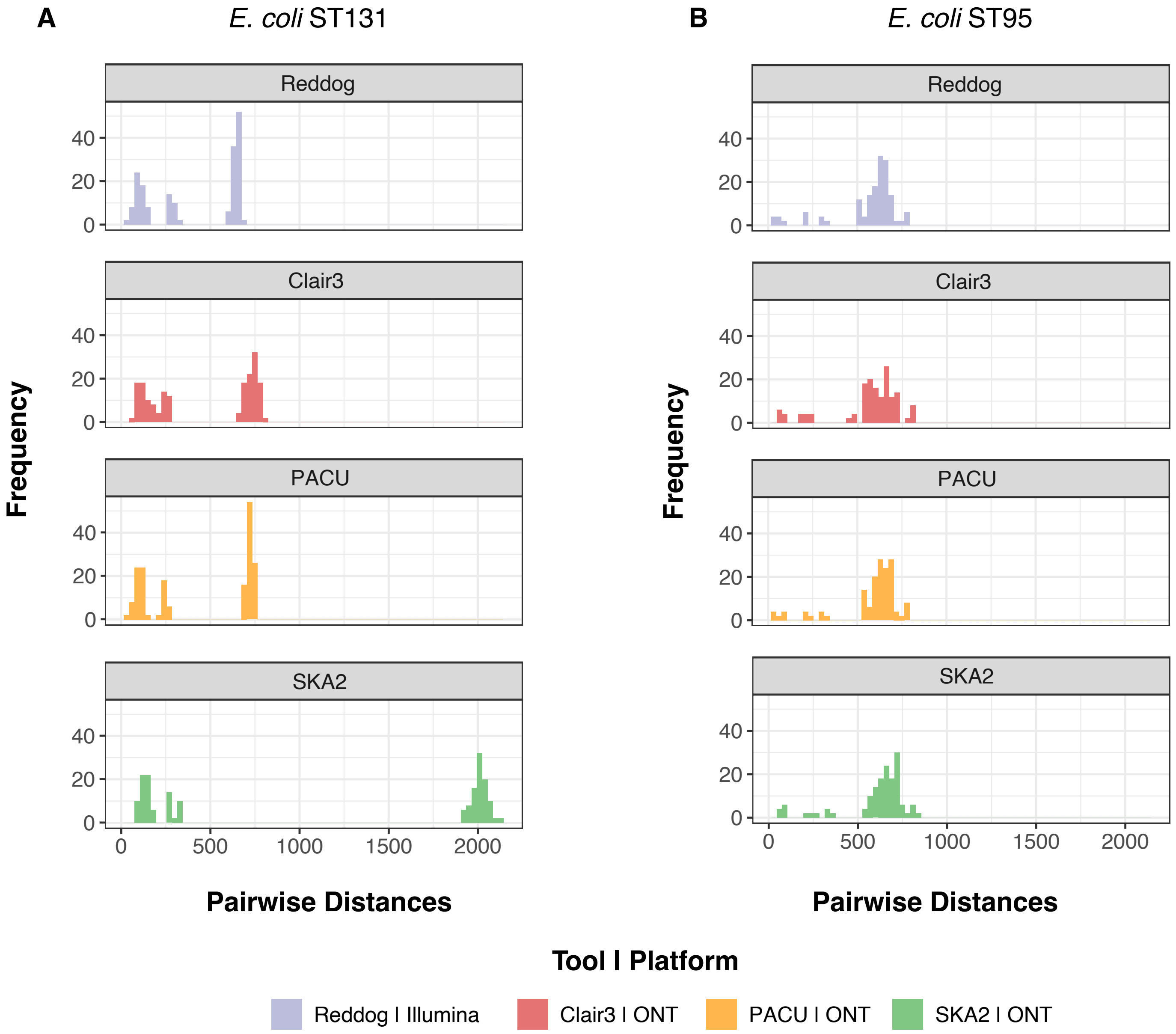


**Supplementary Figure 9 – Pairwise distance distribution of *E. coli* isolates after calling core genome SNPs using varying variant calling tools.** Distributions are coloured by the tool used. **A)** *E. coli* ST131 isolates. **B)** *E. coli* ST95 isolates.

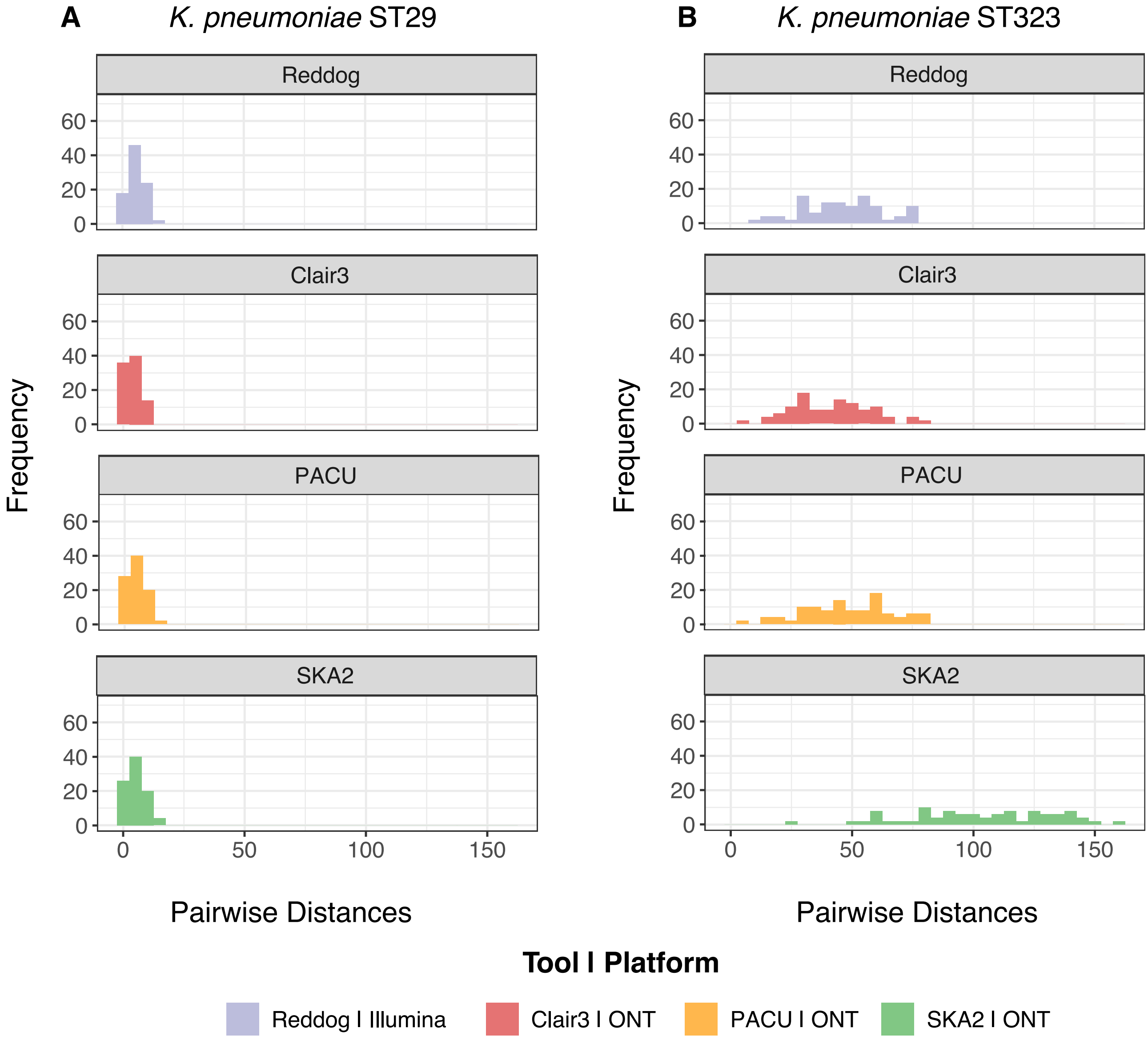


**Supplementary Figure 10 – Pairwise distance distribution of *K. pneumoniae* isolates after calling core genome SNPs using varying variant calling tools.** Distributions are coloured by the tool used. **A)** *K. pneumoniae* ST29 isolates. **B)** *K. pneumoniae* ST323 isolates.

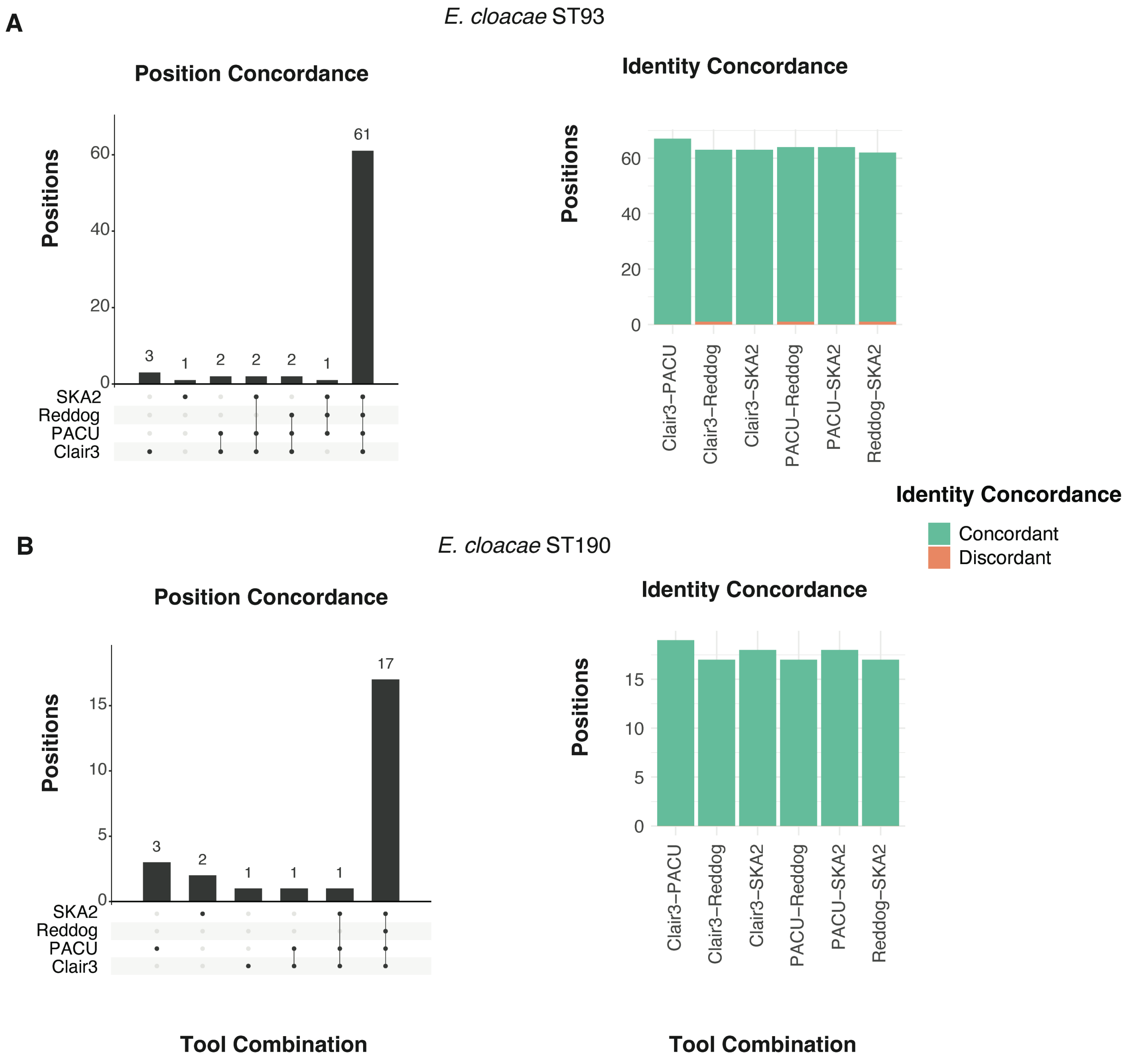


**Supplementary Figure 11 – Concordance between variant calling tools in identifying variant sites and calling a reference or alternative allele in *E. cloacae* isolates.** "Position Concordance" refers to which combination of tools identified a given base position in the reference genome as a variant site. "Identity Concordance" refers to whether all strains were assigned the same allele (either reference or alternative) at positions that were identified as variant sites in both tools. **A)** *E. cloacae* ST93 isolates. **B)** *E. cloacae* ST190 isolates.

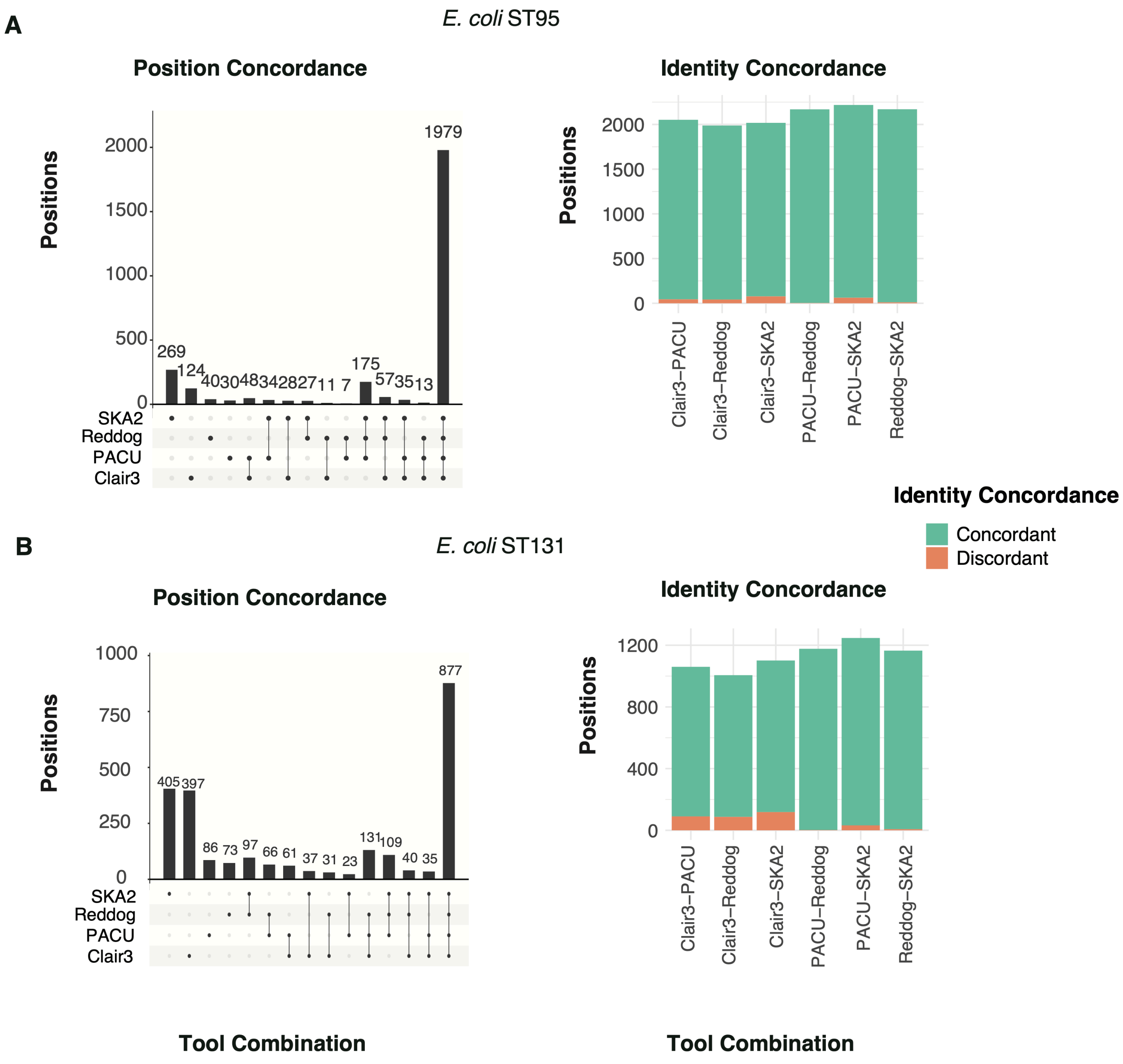


**Supplementary Figure 12 – Concordance between variant calling tools in identifying variant sites and calling a reference or alternative allele in *E. coli* isolates**. "Position Concordance" refers to which combination of tools identified a given base position in the reference genome as a variant site. "Identity Concordance" refers to whether all strains were assigned the same allele (either reference or alternative) at positions that were identified as variant sites in both tools. **A)** *E. coli* ST95 isolates. **B)** *E. coli* ST131 isolates.

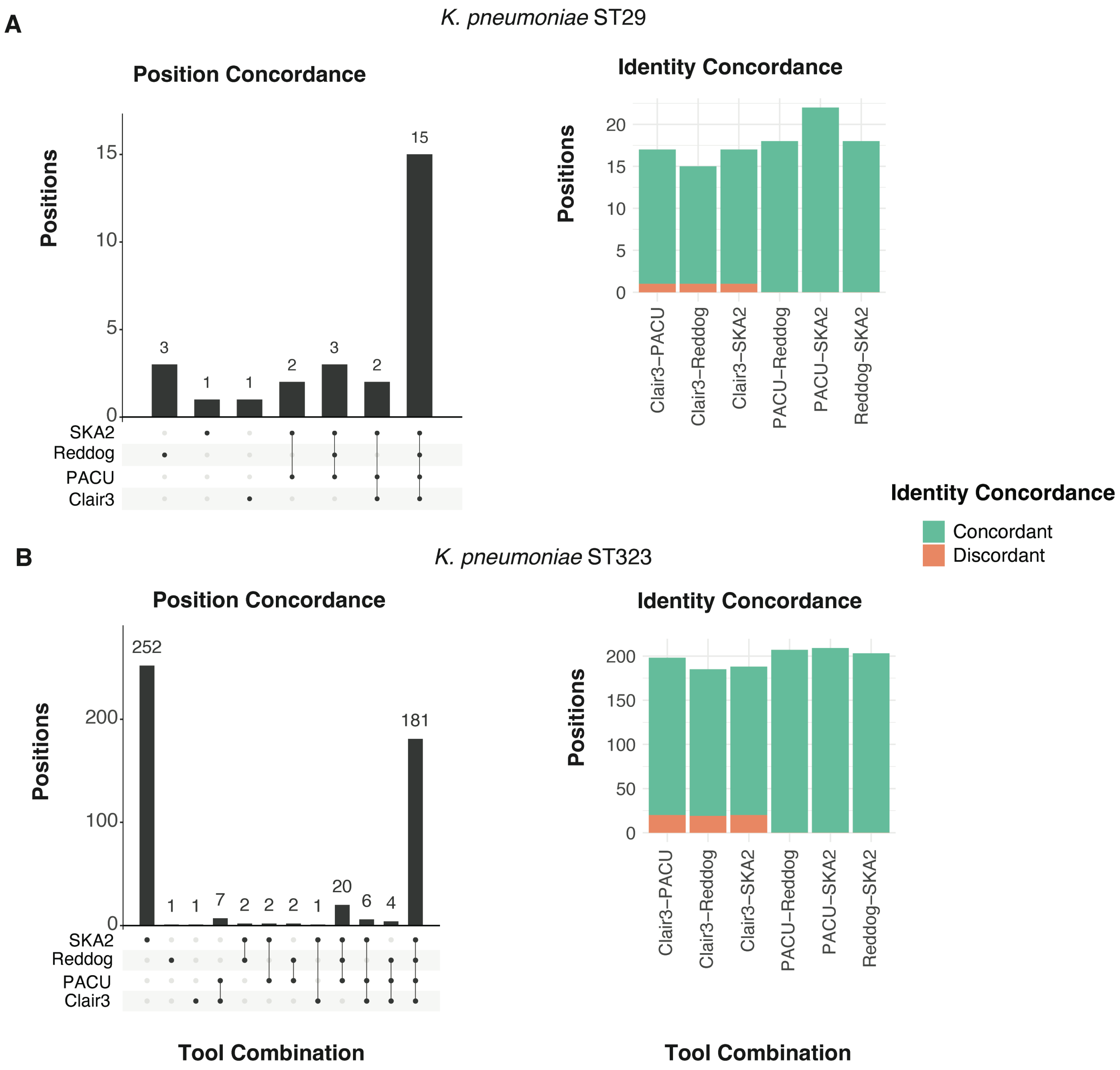


**Supplementary Figure 13 – Concordance between variant calling tools in identifying variant sites and calling a reference or alternative allele in *K. pneumoniae* isolates**. "Position Concordance" refers to which combination of tools identified a given base position in the reference genome as a variant site. "Identity Concordance" refers to whether all strains were assigned the same allele (either reference or alternative) at positions that were identified as variant sites in both tools. **A)** *K. pneumoniae* ST29 isolates. **B)** *K. pneumoniae* ST323 isolates.

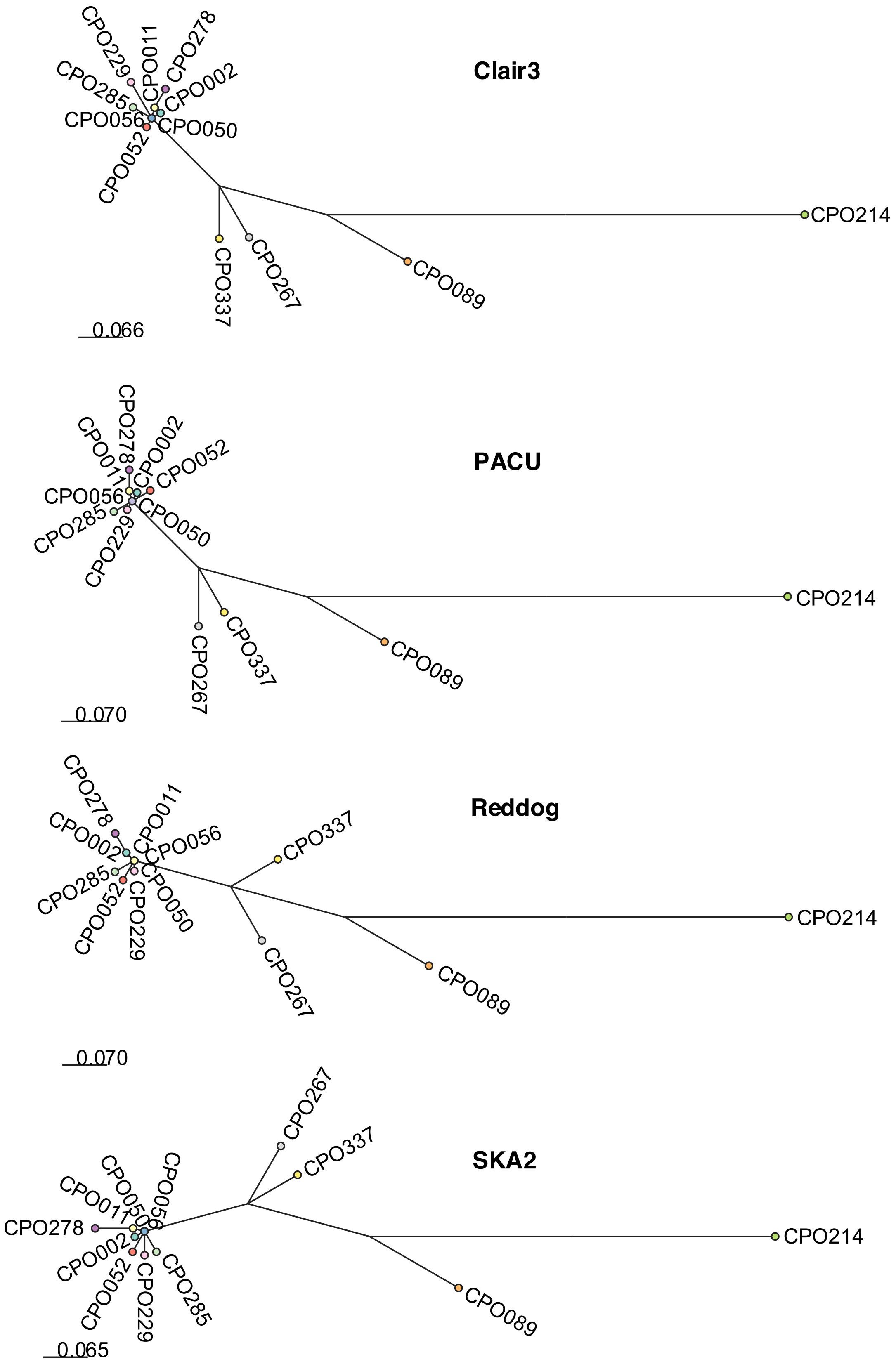


**Supplementary Figure 14 - Concordance between ONT methods and traditional Illumina methods in constructing *E. cloacae* ST95 phylogenies**. nRF refers to Normalised Robinson-Fould distance, which calculates topological similarity (0 = identical, 1 = no shared topology).

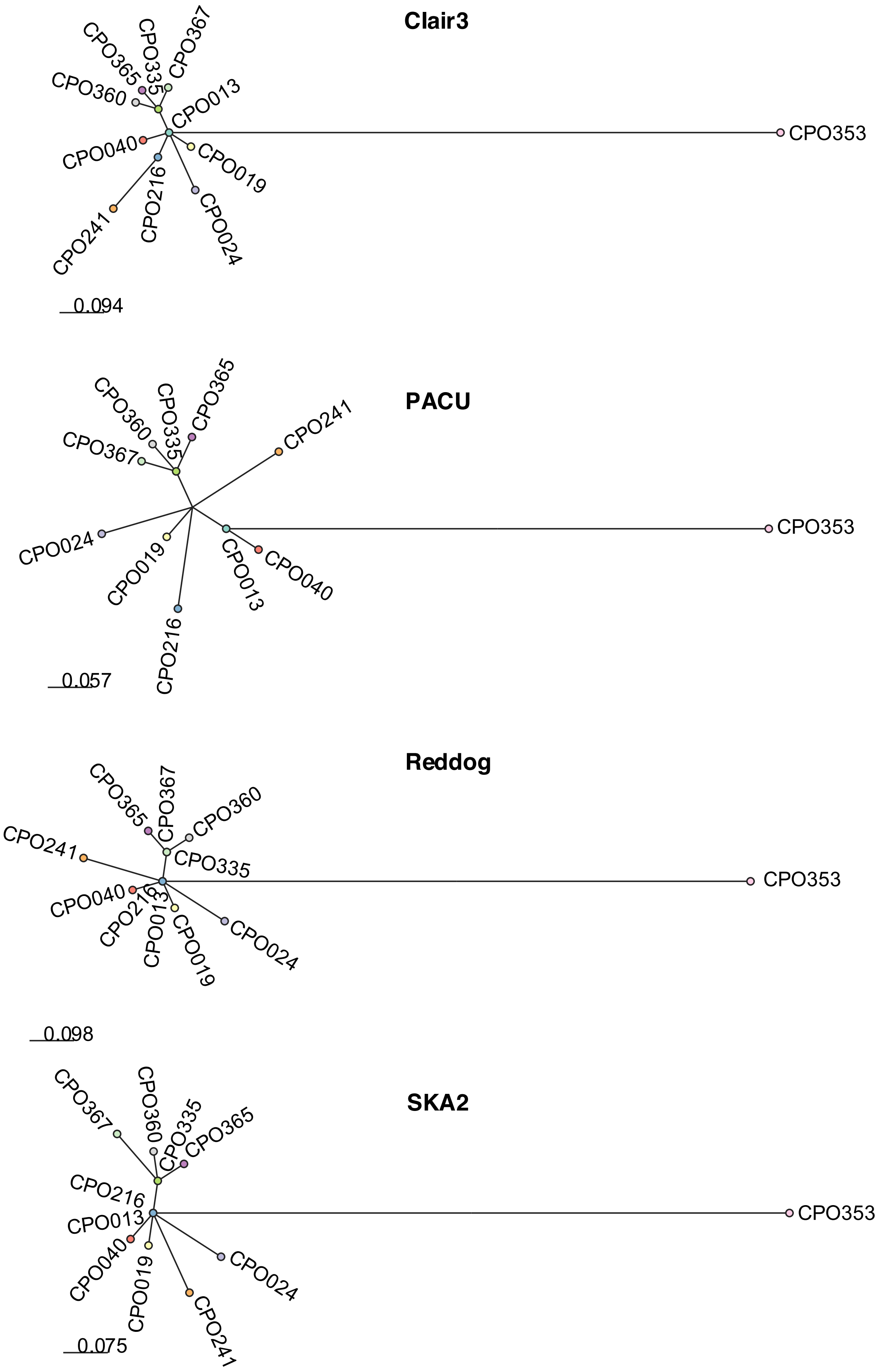


**Supplementary Figure 15 - Concordance between ONT methods and traditional Illumina methods in constructing *E. cloacae* ST190 phylogenies**. nRF refers to Normalised Robinson-Fould distance, which calculates topological similarity (0 = identical, 1 = no shared topology).

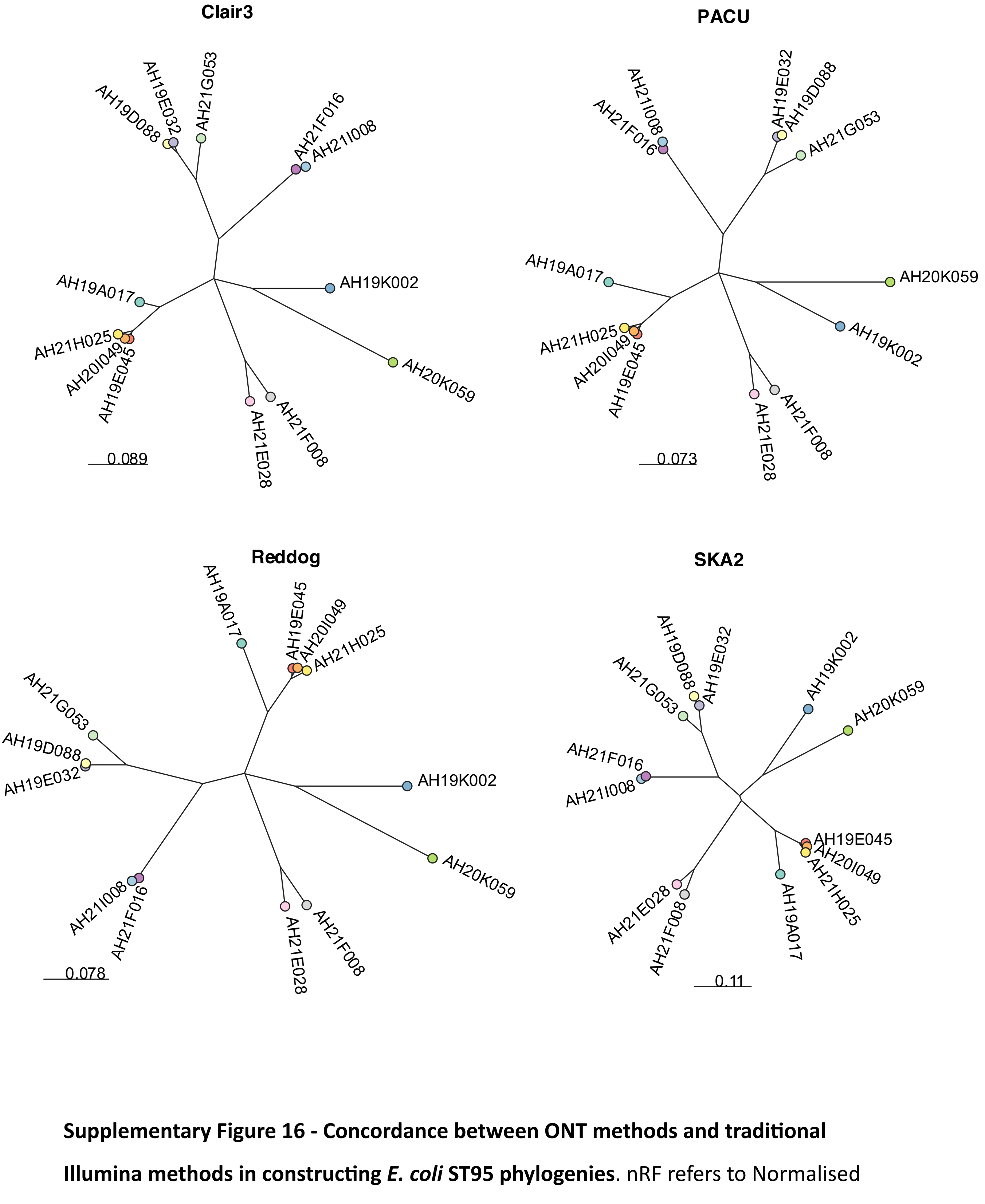


**Supplementary Figure 16 - Concordance between ONT methods and traditional Illumina methods in constructing *E. coli* ST95 phylogenies**. nRF refers to Normalised Robinson-Fould distance, which calculates topological similarity (0 = identical, 1 = no shared topology).

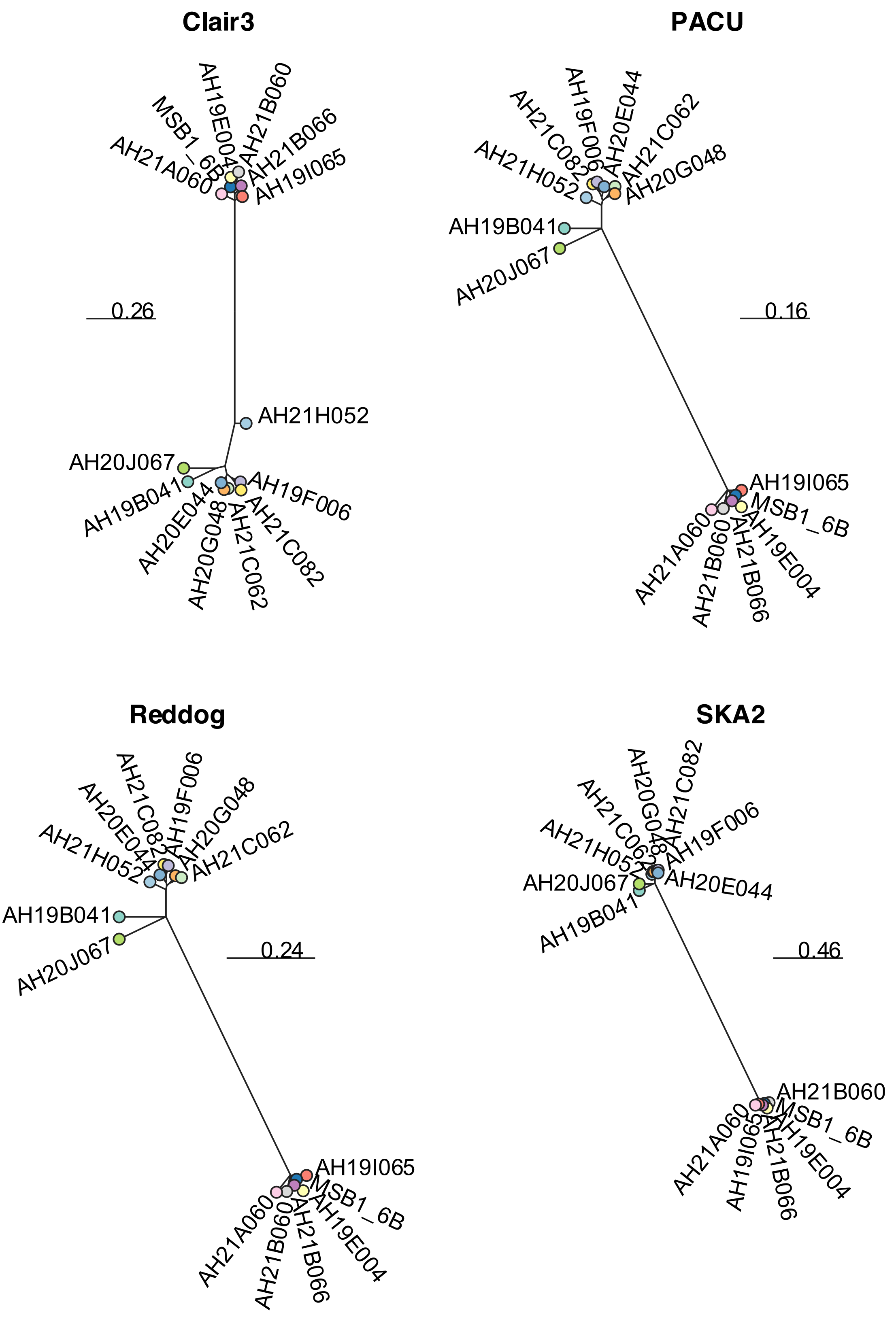


**Supplementary Figure 17 - Concordance between ONT methods and traditional Illumina methods in constructing *E. coli* ST131 phylogenies**. nRF refers to Normalised Robinson-Fould distance, which calculates topological similarity (0 = identical, 1 = no shared topology).

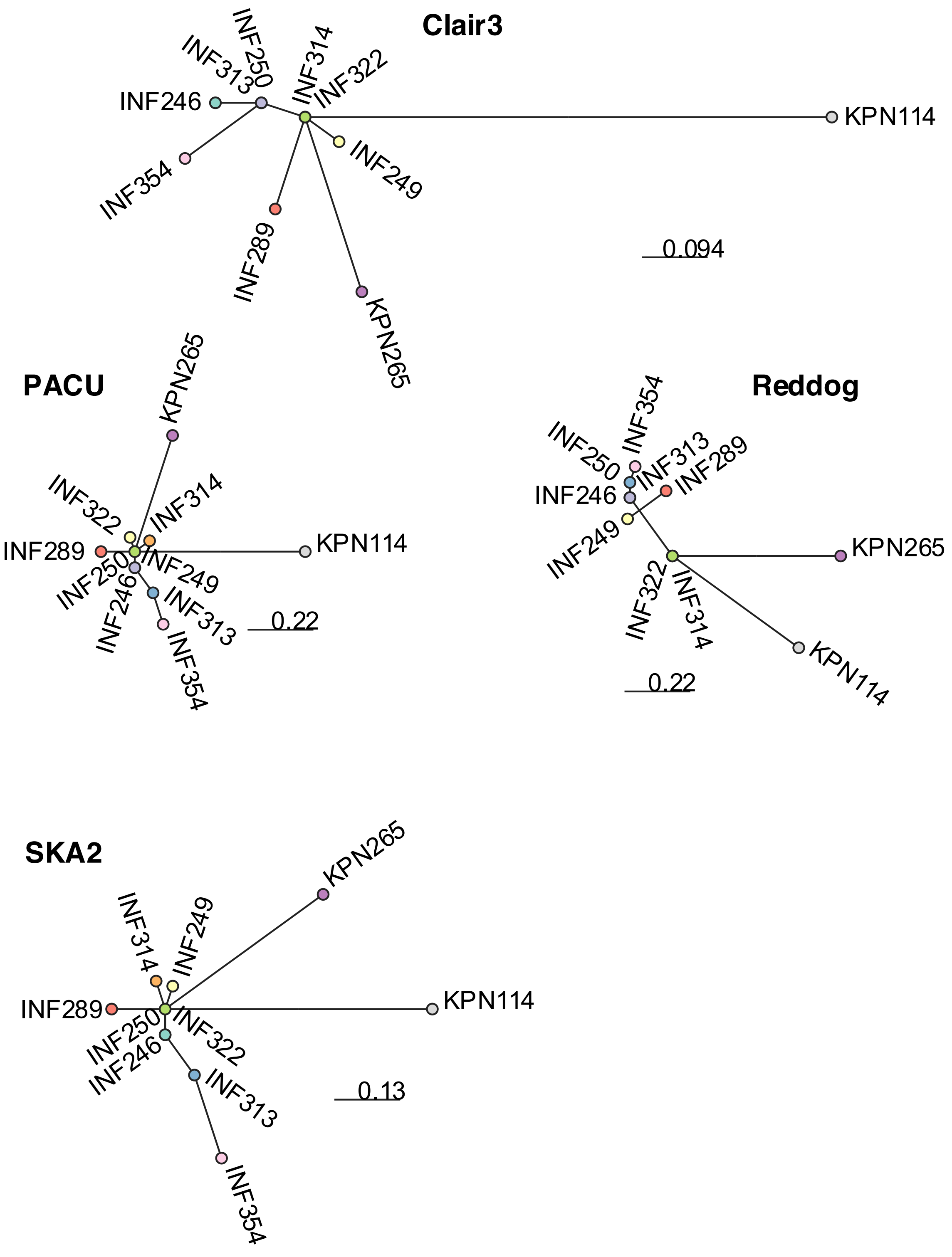


**Supplementary Figure 18 - Concordance between ONT methods and traditional Illumina methods in constructing *K. pneumoniae* ST29 phylogenies**. nRF refers to Normalised Robinson-Fould distance, which calculates topological similarity (0 = identical, 1 = no shared topology).

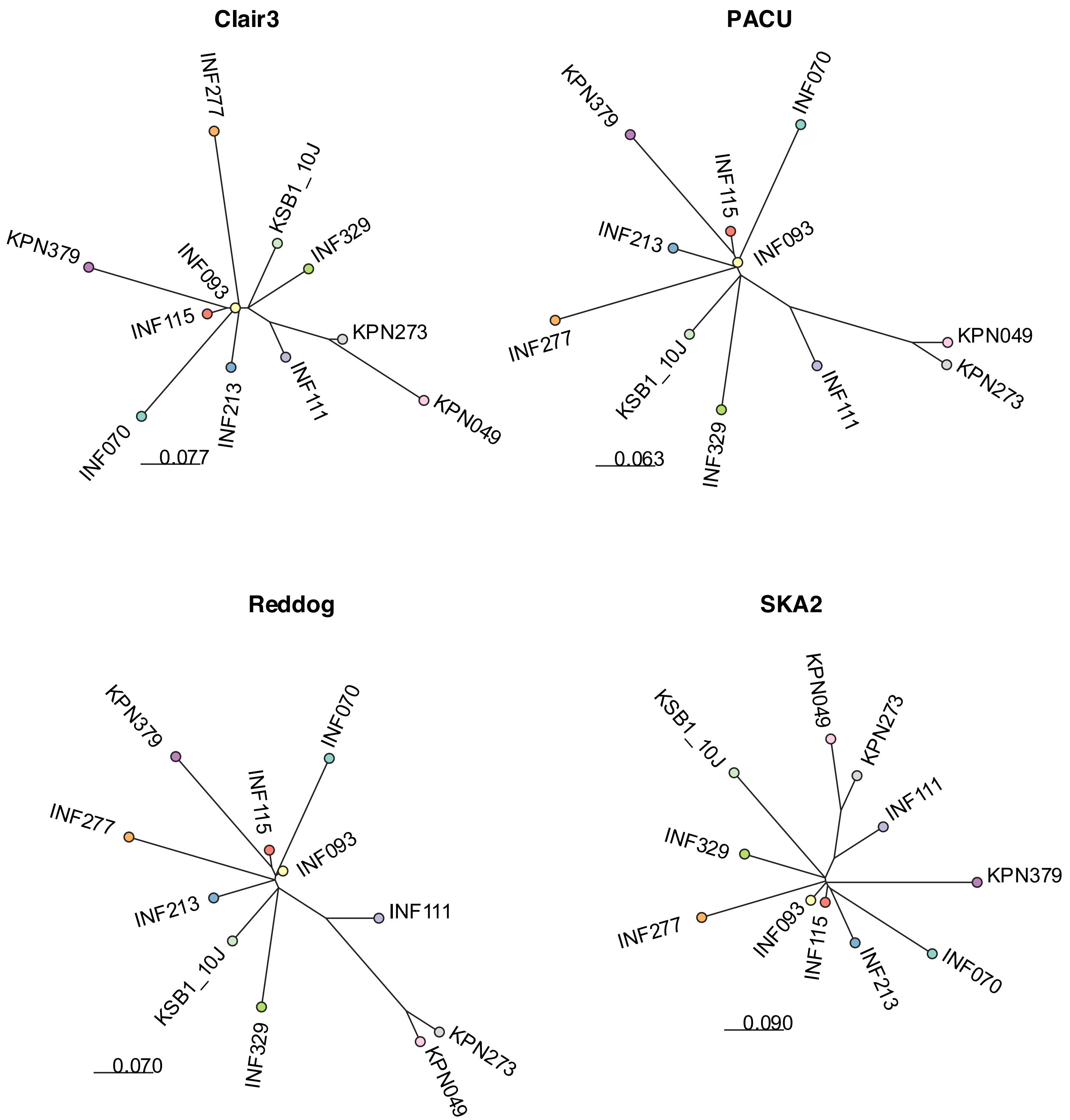


**Supplementary Figure 19 - Concordance between ONT methods and traditional Illumina methods in constructing *K. pneumoniae* ST323 phylogenies**. nRF refers to Normalised Robinson-Fould distance, which calculates topological similarity (0 = identical, 1 = no shared topology).

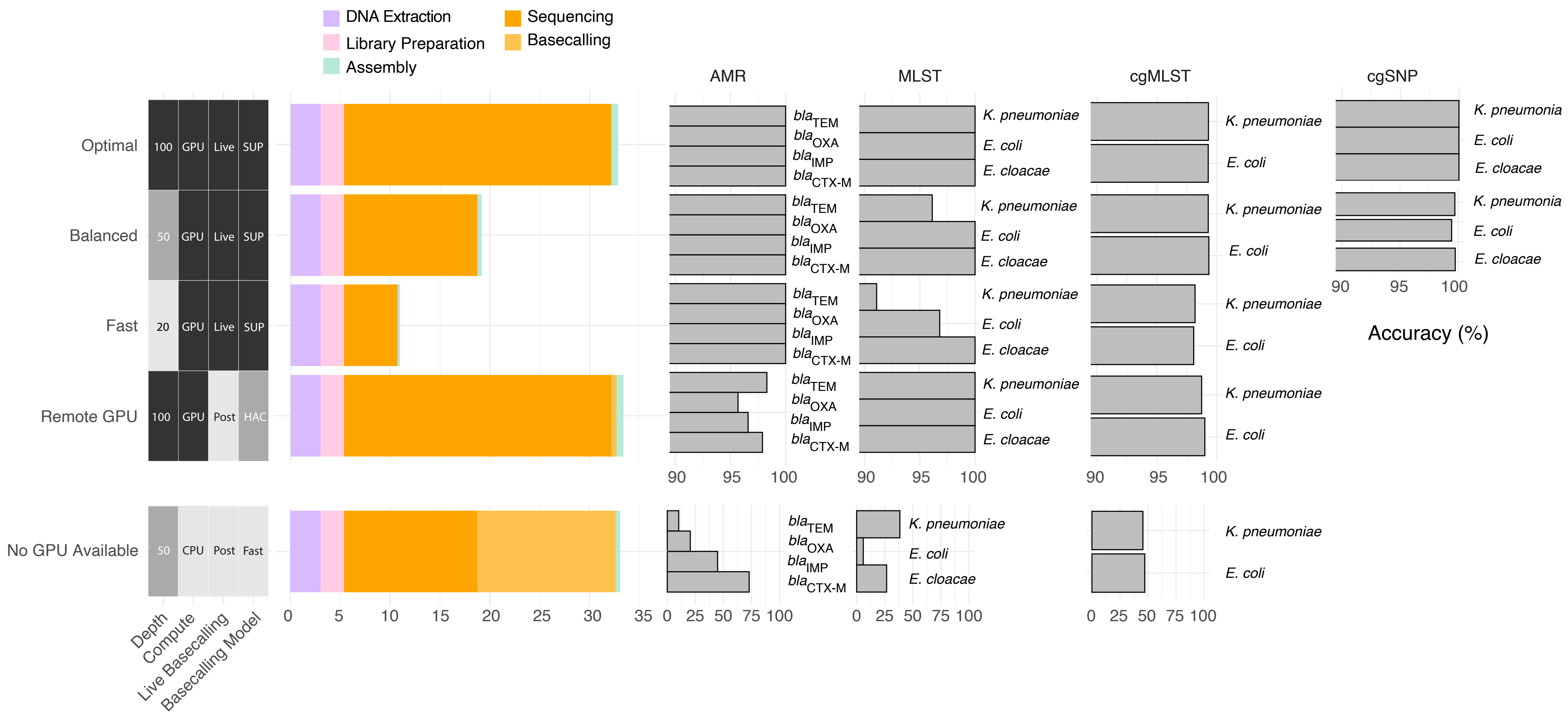


**Supplementary Figure 20 - Time requirements and expected analysis accuracy across five methods for multiplexed ONT sequencing of 20 bacterial isolates.** Accuracy metrics summarise the benchmarking outcomes using R10.4.1 Dorado polished assemblies for MLST, cgMLST and AMR, except for AMR typing at 20x depth where raw reads are recommended. cgSNP accuracy was summarised from simulated R10.4.1 readsets. Accuracy metrics are coloured by GPU availability (blue=available, red=unavailable). DNA extraction and library preparation time requirements are sourced from Genfind and ONT's protocol approximations. Sequencing time was calculated based on all ONT runs that contributed to the benchmarking dataset. Basecalling and assembly time requirements were benchmarked on an NVIDIA A100 GPU with 60 CPUs and 40GB of RAM.
